# Increased plasma fibronectin mirrors intimal phenotypic switching of vascular smooth muscle cells in moyamoya arteriopathy

**DOI:** 10.1101/2025.09.26.25336314

**Authors:** Caroline Asselman, Jozefien Meersschaut, Patrick Willems, Julien Mortier, Frank Vernaillen, Elisabeth Dhondt, Anne Sieben, Sasha Libbrecht, Esperanza Fernandez, Kris Gevaert, Ward De Spiegelaere, Evelien Van Hamme, Veerle De Herdt, Jo Van Dorpe, Dimitri Hemelsoet, Bart Dermaut, Francis Impens

## Abstract

Moyamoya disease (MMD) is a rare cerebrovascular disorder characterized by progressive stenosis of large intracranial arteries and formation of fragile collateral vessels that can be triggered by a broad range of genetic and immune factors. Central to MMD pathology is excessive proliferation of vascular smooth muscle cells (VSMCs) in the intima of affected arteries associated with contractile-to-synthetic phenotypic switching, but the underlying molecular mechanisms remain unclear. To identify dysregulated pathways we studied a cohort of 12 patients with well-documented MMD, including one post-mortem autopsy case, using a multi-omics approach combining whole exome sequencing with plasma proteomics. In addition, we conducted an in-depth spatial proteomics analysis of an occluded artery retrieved post-mortem from one idiopathic patient, combining targeted antibody-based imaging with laser capture microdissection coupled to mass spectrometry. Genetic predispositions for MMD was found in 8 out of 12 patients (67%), including three patients with variants in the major susceptibility gene *RNF213* and five with varying underlying genetic conditions (trisomy 21, pathogenic variants in *ACTA2*, *SAMHD1*, *NFIA*). Artery spatial proteomics revealed phenotypic switching of vascular smooth muscle cells associated with infiltration of these cells in the intima, including loss of contractile and gain of synthetic marker proteins. Most notably, increased expression of cellular fibronectin in the occluded lesion was associated with increased levels in patients’ plasma, providing a rational for cellular fibronectin as potential tissue leakage biomarker for moyamoya disease. Finally, infiltration of macrophages and antigen-presenting cells in the intima pointed to a role for inflammatory signals in disease progression. Together, our data provide an unprecedented spatial view on protein changes in an occluded moyamoya artery, revealing intimal infiltration of synthetic vascular smooth muscle cells and antigen-presenting immune cells as key pathological findings, opening novel avenues for future diagnosis and research.

## Introduction

Moyamoya disease (MMD) is a rare and progressive cerebrovascular disorder characterized by the gradual stenosis and occlusion of intracranial arteries, mainly the distal internal carotid artery and proximal middle and anterior cerebral arteries. Occlusion of these arteries is the result of abnormal infiltration and proliferation of vascular smooth muscle cells (VSMCs) in the intima in response to yet unknown stimuli.^1^ Typical for MMD is the formation of compensatory collateral fragile vessels, angiographically resembling a “puff of smoke” (“moyamoya” in Japanese).^2^ Clinically, MMD is often characterized by transient or permanent ischemic events or hemorrhagic stroke.^3^ Symptoms include focal neurological deficits related to reduced or absent blood flow to the brain such as dysarthria, aphasia, hemiparesis, sensory disturbances, visual impairments, involuntary movements, seizures, altered consciousness, and cognitive dysfunction. Additionally, non-specific symptoms like vertigo, dizziness, or headaches may occur due to cerebral hemodynamic compromise or meningeal collateral vessel dilation.^3^ When these vascular changes, also referred to as moyamoya angiopathy (MMA), occur in association with another condition the term moyamoya syndrome (MMS) is frequently used.^3,4^

The precise etiology of MMD is unknown, however, genetic factors have long been suspected to play a crucial role. In 9-15% of patients, a positive family history of MMD is observed.^5^ In addition, the association of MMD or MMA-like changes with multiple genetically transmitted disorders such as trisomy 21, neurofibromatosis type 1 or pathogenic variants in *ACTA2* and *SAMHD1,* as well as the occurrence of a strong ethnic preference in East Asia suggest an important genetic component.^4^ In 2011, a missense variant within the *RNF213* gene known as the p.(R4810K) founder variant was found to dramatically increase MMD risk by over 100-fold in East-Asian populations.^6,7^ While homozygous carriers of this variant have a ∼80% chance of developing MMD, heterozygous carriers show a much lower disease penetrance, suggesting that beyond genetic predisposition, a second environmental and/or genetic ‘hit’ is needed to trigger MMD onset.^4^ Interestingly, recent molecular studies uncovered RNF213 as a multifunctional protein with a pivotal role in the innate immune response against microbial infections.^4,8–10^ In addition, a growing body of clinical evidence shows that MMD often occurs in conjunction with infection or autoimmune conditions. Together, these molecular and clinical observations suggest a role for immune-related triggers as a second hit in moyamoya pathogenesis in genetically predisposed individuals.^4^

While the *RNF213* p.(R4810K) founder variant largely explains the increased MMD prevalence in East Asia, other *RNF213* variants and other genes have also been reported in association with MMD in and outside the East Asian population. However, genome-wide association studies have yet to pinpoint predominant susceptibility variants in Caucasian MMD.^11^ Since comparatively limited research has been conducted on patients with MMA in Western countries,^12–15^ we clinically and genetically characterized a cohort of 12 patients with MMA followed at the University Hospital in Ghent, Belgium, and profiled their plasma proteome in comparison with healthy individuals. In addition, a detailed histopathological and single-cell spatial proteomics analysis of an occluded artery of one of the patients was performed, providing unprecedented insight into VSCM phenotypic switching and immune cell infiltration in MMD development. Upregulation of cellular fibronectin in the occluded artery correlated with increased plasma levels of cellular fibronectin, which we propose here as a potential biomarker for MMD.

## Material and methods

### Patient cohort

From 2019 until 2022 we recruited a cohort of 12 affected individuals with MMA at Ghent University Hospital (Universitair Ziekenhuis Gent or UZ Gent). All patients were clinically evaluated, including medical and family history, and diagnostic imaging. Cerebral computed tomographic imaging, magnetic resonance imaging, and conventional digital subtraction angiography were performed in these patients at multiple timepoints. MMD/MMS diagnosis was made according to international guidelines.^16^ Blood samples of these 12 patients were taken for multiomics work-up including whole exome sequencing (WES) and plasma proteomics. Formalin-fixed paraffin-embedded (FFPE) brain autopsy material was generated from one MMD patient and used for spatial omics analysis of an occluded MMD artery.

### Genetic studies

Genetic studies were initiated at the Center for Medical Genetics at UZ Gent and included WES of all patients except patient 11 for whom WES had been performed previously in a research setting.^18^ Molecular diagnoses had already been obtained in four more patients during diagnostic work-up prior to the start of this study using different genetic techniques: patient 2 (conventional karyotyping), patient 3 (molecular karyotyping by array CGH and shallow whole genome sequencing) and patients 4 and 8 (targeted gene analysis). WES was performed on the Illumina Novaseq 6000 Platform after enrichment of gDNA with SureSelectXT Low Input Human All Exon v6 and v7 (Agilent Technologies). The BWA-MEM 0.7.17 algorithm was used for read mapping against the human genome reference sequence (NCBI, GRCh38/hg38) duplicate read removal, and variant calling. Variant calling and filtering were performed using Seqplorer, an in-house developed tool for the analysis of ES data. The position of the called variants was based on NCBI build GRCh38. A minimum of 90% of the interrogated genes had a coverage of >20x. Nucleotide numbering was according to the Human Genome Variation Society guidelines (HGVS). Variant filtering criteria in Seqplorer included a population frequency (gnomAD) <0.02, impact on the protein predicted to be moderate or high, minimal variant quality of 20, and minimal depth of 2 reads. Low-impact variants such as synonymous variants or intronic splice region variants >8 nucleotides away from the splice junction were not prioritized using standard settings. Potential CNVs were called using ExomeDepth, an algorithm which uses WES data to detect read depth differences in coding regions.^17^

Analysis of candidate genes was performed in all patients in two ways: (1) the application of a small Online Mendelian Inheritance in Man (OMIM)-based candidate gene list of 18 genes previously reported in MMD/MMS-related monogenic disorders (**Table 1**) and (2) a complete Mendeliome analysis (4878 OMIM genes). Variant classification was performed using an in-house developed tool based on the ACMG and ACGS guidelines^18^ in the following classes: (1) benign, (2) likely benign (>95% certainty that the variant is benign), (3) variant of unknown significance, (4) likely pathogenic (>95% certainty that the variant is pathogenic), and (5) pathogenic.^18^

**Table 1.**
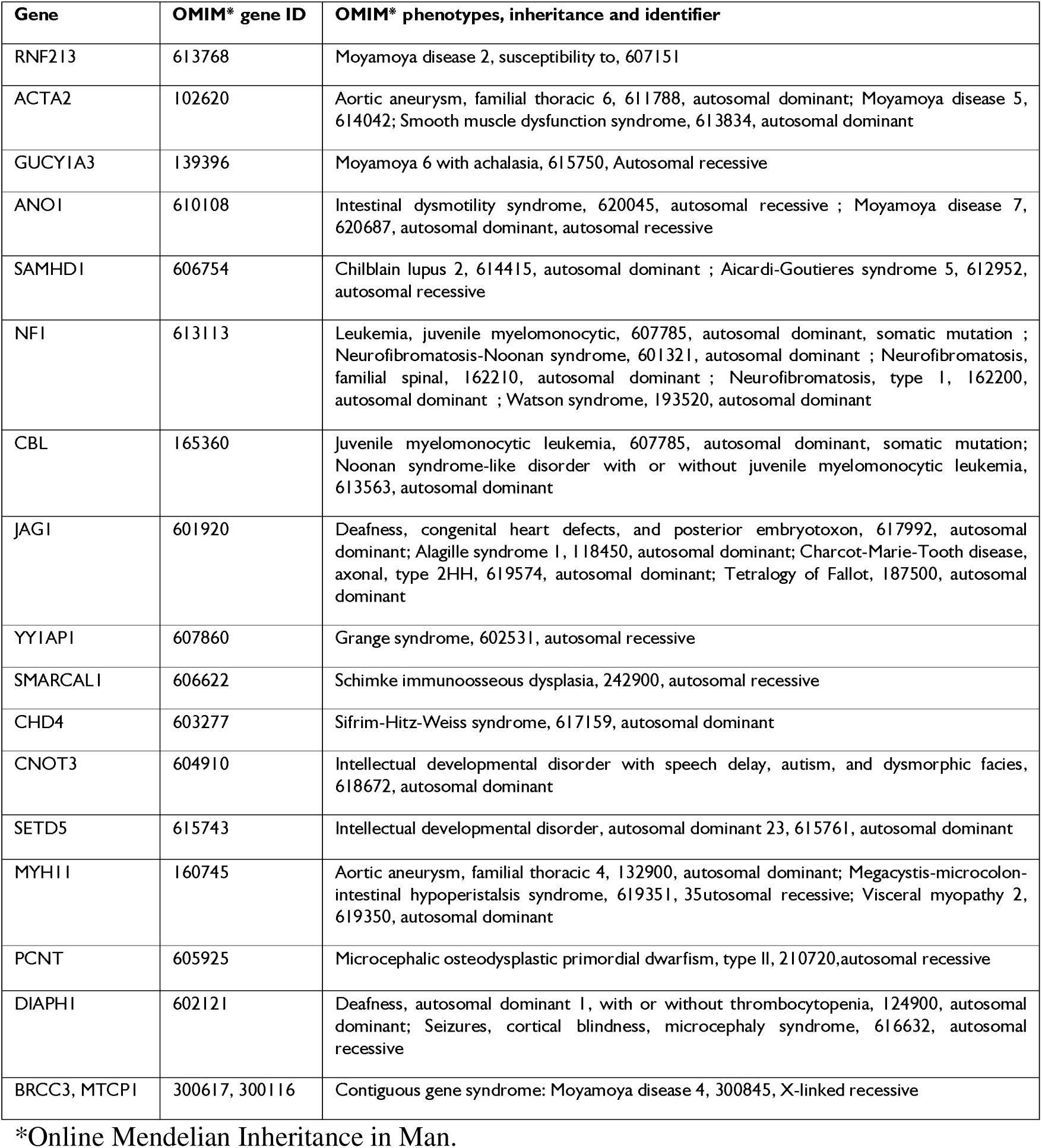
Diagnostic OMIM candidate gene list for moyamoya disease/moyamoya syndrome.

### Brain sample immunohistochemistry and laser capture microdissection

#### Processing and preservation of brain sample

Tissue samples of the autopsy case were provided by the pathology department of UZ Gent. Autopsy of the brain pathology was performed according to the standardized protocols of UZ Gent. Dissection of the large brain arteries and the circle of Willis at the base of the brain was performed on fresh brain tissue. Next, one hemisphere was sliced and frozen at -80°C, and the other hemisphere was fixed in neutral buffered 4% formaldehyde, and processed to paraffin embedding with an automatic processor (Microm STP 420D, Thermo Fisher Scientific, Massachusetts, USA).

#### Immunohistochemistry

Paraffin blocks of transverse sections of the large brain arteries and circle of Willis were sectioned into 4 μm thick histological sections and stained with hematoxylin and eosin (H&E). Immunohistochemical (IHC) stainings for smooth muscle actin, calponin, H-caldesmon, desmin, CD45, CD3, CD8, CD20, CD4, CD68, ERG and CD31 were performed in routine fashion using a Ventana immunostainer (Roche, Basel, Switzerland).

#### Laser capture microdissection

Paraffin-embedded tissues were cut into 8 μm thick sections and mounted on PEN membrane slides (ZEISS, Oberkochen, Germany). The slides were left in an incubator at 56°C overnight for optimal adherence of the tissue to the membrane. Slides were dewaxed, rehydrated, and then stained in hematoxylin (Merck KGaA, Darmstadt, Germany) for 10 s, followed by rinsing in water (10 s) and Scott’s tap water solution (10 s) (Sigma-Aldrich, Diegem, Belgium). Subsequently, the tissue slides were stained with eosin Y (Merck KGaA, Darmstadt, Germany) for 5 s and dehydrated in graded ethanol concentrations, starting with two baths of 95% for 10 s followed by two baths of isopropanol for 30 s. Finally, the sections were completely dehydrated by immersing the slides in two successive baths of xylene for 60 s each. The slides were air-dried in a laminar flow cabinet and individually placed in sterile 50 ml falcon tubes to prevent rehydration by air. A PALM Microbeam II (Zeiss, Oberkochen, Germany) was used for laser microdissection. Intimal and medial tissue was microdissected and collected in 200 µl adhesive caps.

### Spatial proteomics

For plasma proteomics and laser capture microdissection proteomics analysis methods please see Supplemental Material.

#### Sample preparation and data acquisition

Spatial proteomics analysis was performed using the MACSima™ Imaging System, a fully automated instrument that combines widefield microscopy and liquid handling for cyclic immunofluorescence imaging. Staining cycles consisted of the following automated steps: immunofluorescent staining, sample washing, multi-field imaging, and signal erasure (photobleaching or REAlease). FFPE brain tissue was sectioned on slides and a MACSWell^TM^ imaging frame was mounted immediately on the slide. Ice-cold 4% PFA solution was added according to the MACSWell^TM^ imaging frames datasheet and incubated for 10 min at room temperature. The slide was washed three times with MACSima Running Buffer. After washing, the initial sample volume of MACSima Running Buffer was added according to the MACSWell^TM^ imaging frames datasheet. Right before the start of the MACSima^TM^ instrument a DAPI pre-staining was performed. To this end, the MACSima Running Buffer was removed from the sample to be analyzed and stained for 10 min with a 1:10 dilution of a DAPI staining solution. The DAPI staining solution was removed and three washing steps were performed with MACSima Running Buffer. Finally, the initial sample volume of MACSima Running Buffer was added. We used the REAscreen MAX kit, human, FFPE, version 02 (Miltenyi Biotech, Bergisch Gladbach, Germany), in addition to six antibodies against human ISG15, RNF213, MMP2, FN1, IFN-γ and ACTA2 (**Supplementary Table 1**).

#### Image analysis

The raw data outputted by the MACSima^TM^ device was preprocessed using the accompanying software suite, MACS IQ View Analysis. Among other imaging corrections (e.g. flatfield correction), this preprocessing step performs stitching and registration based on the DAPI-images in each imaging cycle. Moreover, for each cycle background correction was performed on the biomarker signal images by subtracting the images acquired after signal erasure of the previous cycle. Of the initial 146 markers from the REAscreen MAX panel, 15 (Cytokeratin, CytokeratinHMW, BATF, TIM3, MastCellTryptase, CD209, MelanocytePMEL, CLA, Dnmt3b, PKCalpha, Cytokeratin14, CD66, CD202b, CD100 and CD134) were removed due to clear quality issues. Using custom Python scripts, the resulting TIFF images of the imaged region of interest (ROI) were cropped to focus on a high quality ROI devoid of major acquisition artifacts (i.e. focusing issues, tissue folds, etc.) and for some channels, the automatic registration from the MACS IQ View was manually corrected by including an extra 15-pixel shift. For the first three cycles (containing the six custom antibodies), a larger correction in registration was needed, which was achieved by placing manual landmarks on the DAPI-image of cycle 1 and the DAPI-image of cycle 4 and transforming the images of the first three cycles using a thin-plate spline transformation in the BigWarp plugin in FIJI (ImageJ 1.54f).^19^ Next, tissue regions (lumen, intima, etc.) and artifacts still present in the cropped ROI (out-of-focus, tissue folds and acquisition bleaching) were manually annotated in QuPath (v.0.4.4)^20^ to remove any cells detected in these regions. Using the python package SPArrOW as an analysis framework, nucleus segmentation was performed by Cellpose^21^ on the DAPI-channel. Cellpose was also used to estimate a whole-cell segmentation for immune cells based on a combination of immune markers (CD15, CD43, CD45, CD45RA, CD45RB, CD57, CD66b, CD162, HLADR and Syk) that had good signal-to-noise and where the cell outlines could be detected. Whole-cell segmentation for red blood cells was based on the CD233 marker. For all other cells, the cell outline was approximated by expanding the nucleus segmentation by 10 pixels. Using these cell segmentations, object classifiers were trained for marker positivity in ilastik (v.1.4.0.post1)^22^ for CD2, CD8a, CD15, CD43, CD45, CD45RA, CD45RB, CD57, CD66b, CD162, HLADR and Syk based on the intensity features of the corresponding marker image.

#### Single cell analysis

Average intensities per cell for all channels were used to generate a 10X-like input containing 138 features (antibody channels) for 3,936 cells to enable single cell analysis with Seurat (version 4.3.0.1).^23^ A Seurat object was generated using the Read10X function and cell metadata from the spatial proteomics image analysis was added, including the manually annotated tissue region (e.g. intima or tunica media), the presence of technical artefacts and others. Cells presenting outlier intensity values, acquisition bleaching or residing in out-of-focus or tissue folding areas were removed. For data normalization, we performed centered log ratio (CLR) transformation within cells (NormalizeData function, method ‘CLR’, margin 2). Afterwards, we detected highly variable genes (FindVariableGenes function), scaled data (ScaleData function) and performed principal component analysis (runPCA function) using 12 principal components. Subsequently, unsupervised clustering was performed using the FindNeighbours and FindClusters functions (clustering resolution of 0.2) and visualized by uniform manifold approximation and projection (UMAP) using the RunUMAP function. This reveals five clusters corresponding to distinct cell tissues (intima, adventitia, media, endothelium and lumen). Protein markers for each cluster were identified with the FindMarkers function using the ‘roc’ test (upregulated markers only), where we retained the top five ranked proteins with the highest predictive power for each cluster. This procedure was repeated for the subset of 223 immune cells identified by the whole-cell segmentation based on a combination of immune markers described above.

#### Data availability

The mass spectrometry proteomics data of the laser capture microdissection (LCM) and plasma proteomics have been deposited to the ProteomeXchange Consortium (http://proteomecentral.proteomexchange.org) via the PRIDE partner repository^24^ with the dataset identifiers PXD057183 and PXD063236, respectively.

## Results

### Patient cohort: clinical and genetic characteristics

Twelve patients, diagnosed with MMD or MMS were included. The study cohort consisted of ten unrelated sporadic cases and one hereditary MMD. A summary of the clinical and genetic characteristics of all patients is shown in **Table 2**, comprehensive patient information is provided as **Supplementary Material**. Blood samples were obtained from all twelve patients. The brain of one patient was obtained for further investigation. Nine patients showed bilateral MMD versus unilateral MMD in three patients. All patients had experienced at least one ischemic (nine patients) or hemorrhagic stroke (three patients). Three patients received neurosurgical treatment with revascularization. The most common symptoms reported were migraine without aura (three patients), arterial hypertension (four patients), and recurring viral infections of the upper airway (five patients). Interestingly, five out of twelve patients were diagnosed with other conditions in addition to MMD, such as Down syndrome, Crohn’s disease, diabetes mellitus type 2 and cancer (osteosarcoma in the jaw).

**Table 2.** Clinical and genetic characteristics of all patients.

|  | Age range (years) | Medical history | Clinical presentation | Infection / inflammation | MMA uni- or bilateral / collaterals | Genetic diagnosis | ClinVar<sup>†</sup> / gnomAD<sup>‡</sup> / DGV<sup>‡</sup> Frequency |
| --- | --- | --- | --- | --- | --- | --- | --- |
| 1 | 31-35 | No major problems | Collaps with stupor and right hemiparesis | Aspiration pneumonia & cutaneous herpes simplex in acute phase | Bilateral / yes | *RNF213* c.14576G>A p.R4810K,<sup>6</sup> class 5 | Conflicting / 0.000295 |
| 2 | 26-30 | Trisomy 21 | Ischaemic stroke with left hemiparesis | Mycoplasma pneumoniae infection | Unilateral / yes | Trisomy 21<sup>61</sup> | - |
| 3 | 31-35 | Macrocephaly, mild cognitive impairment, pes planus | Ischaemic stroke and partial status epilepticus | Tonsillectomy | Bilateral / yes | *NFIA* intragenic deletion (exon 3-6)<sup>62</sup> | - |
| 4 | 26-30 | Persistent open ductus arteriosus with closure | Neurological evaluation because of delayed motor development; brain-MRI: periventricular leuko-encephalopathy. Cardiac echo: mild dilatation aorta ascendens. | Protracted bronchitis during childhood followed by ataxia, problems with fine motor skills, dysarthria, and involuntary movements with tentative diagnosis of relapsing-remitting Sydenham's chorea | Bilateral / yes | *ACTA2* c.536G>A p.R179H,<sup>30</sup> class 5 | Pathogenic / Not found |
| 5 | 41-45 | Inflammatory bowel disease (Crohn's disease) | Ischemic stroke | folliculitis, Clostridium difficile enteritis | Bilateral / yes |  |  |
| 6 | 61-65 | Headache/migraine since adolescence. Myoma uteri with hysterectomy; arterial hypertension, resection benign lip angioma. | Ischemic stroke with hemiparesis and recurrent TIA | No | Bilateral / yes | *RNF213* g.80364462C>T p.A3927V<sup>29</sup> | - / Not found |
| 7 | 81-85 | Inflammatory arthropathy, spinocellular carcinoma oral cavity, coiling MCA aneurysm | MCA aneurysm | No | Unilateral / yes | *RNF213* g.80364462C>T p.A3927V<sup>29</sup> | - / Not found |
| 8 | 26-30 | Surgery for patent ductus arteriosus with cardiac decompensation at young age, complicated with RSV bronchiolitis ; mydriasis | Fatigue, sensory changes, mydriasis | RSV bronchiolitis at young age | Bilateral / yes | *ACTA2* c.535C>A p.R179S,<sup>30</sup> class 5 | Pathogenic / Not found |
| 9 | 31-35 | Migraine without aura since childhood | Intracerebral haemorrhage | No | Bilateral / yes |  | - |
| 10 | 56-60 | Reumatoid arthritis, migraine | Recurrent ischemic stroke | Chronic sinusitis, acute cholecystitis | Bilateral / yes |  | - |
| 11 | 36-40 | Recurrent mitral valve chorda rupture, type I interferonopathy, short stature | Migraine, pain in the legs during childhood, TIA | Type I interferonopathy | Bilateral / yes | *SAMHD1*<br>c. 676C>T<br>p.R226C,<sup>31</sup> class 3<br>+<br>c. 1608+2 T>C<br>(Splice site mutation),<sup>31</sup> class 4 | Uncertain significance / 0.0000132<br>Likely pathogenic / Not found |
| 12 | 21-25 | RSV bronchiolitis and synovitis hip at young age | Ischemic stroke at young age | RSV bronchiolitis, synovitis | Bilateral / yes |  |  |

†Clinical variant database. ‡Genome aggregation database. \*Database of genomic variants.

*RNF213* variants were found in three MMD patients, including one patient carrying the East-Asian founder variant p.(R4810K) (patient 1).^6,7^ Patients 6 and 7carried another novel rare variant p.(A3927V) of *RNF213*. Similar as p.(R4810K), this mutation is located in the E3 module of RNF213.^25^ In addition, five MMS patients had underlying genetic conditions with known moyamoya associations including trisomy 21 (patient 2) as well as monogenic disorders caused by heterozygous *de novo* pathogenic variants in *ACTA2* (patients 4 and 8),^26^ compound heterozygous variants in *SAMHD1* (patient 11, previously reported)^27^ and a familial heterozygous intragenic deletion in *NFIA* (patient 2). Genetic analysis revealed no known MMD/MMS-related variants in patients 5, 9, 10, and 12.

### Identification of FN1 as a potential biomarker for moyamoya disease

Having established a genetic diagnosis for the majority of our moyamoya patients, we profiled their plasma proteome by untargeted LC-MS/MS analysis to look for immune-related signatures in line with the hypothesis that MMD might be triggered by aberrant immune responses in genetically predisposed individuals.^4^ In addition, quantitative comparison with plasma of sex- and age-matched healthy controls could reveal potential protein biomarkers associated with MMD. Principal component analysis showed coherent clustering of patient and control samples (**Fig. 1A**) and further statistical analysis revealed significantly regulated proteins between both groups **(Fig. 1B**). The control group was characterized by consistently higher levels of platelet marker proteins (**Fig. 1B-C, Supplementary Fig. 1**). These are common contaminations in plasma proteomics data resulting from platelet activation during blood collection.^28^ The absence of these proteins in the MMD group likely results from antiplatelet therapy to which moyamoya patients are subjected as standard of care.^29^ Conversely, we detected six proteins that were significantly upregulated in the moyamoya patient population (**Fig. 1B**), related to blood hemostasis (VWF),^30^ immunity (CRP, C4A),^31,32^ and cell proliferation (OIT3, FBLN5 and FN1).^33–35^ While upregulation of CRP corroborated earlier observations^36,37^ in line with a state of low-grade inflammation in MMD,^38,39^ upregulation of FN1 seemed promising in terms of biomarker potential. Of this protein, two distinct isoforms – plasma Fibronectin (pFN1) and cellular Fibronectin (cFN1) - are described. In contrast to pFN1, cFN1 contains two additional protein domains: extra domain A (EDA) and extra domain B (EDB)^40^ (**Fig. 1D**). pFN1 is produced and secreted by hepatocytes and is abundantly present in plasma, while cFN1 is produced by a broad variety of cell types acting as an extracellular matrix (ECM) protein, with a markedly lower plasma concentration. Interestingly, along with other FN1 peptides, we found an EDA-specific peptide upregulated in moyamoya patients’ plasma, suggesting tissue leakage of cFN1 that we propose here as a potential biomarker for MMD (**Fig. 1D**).

**Figure 1.**
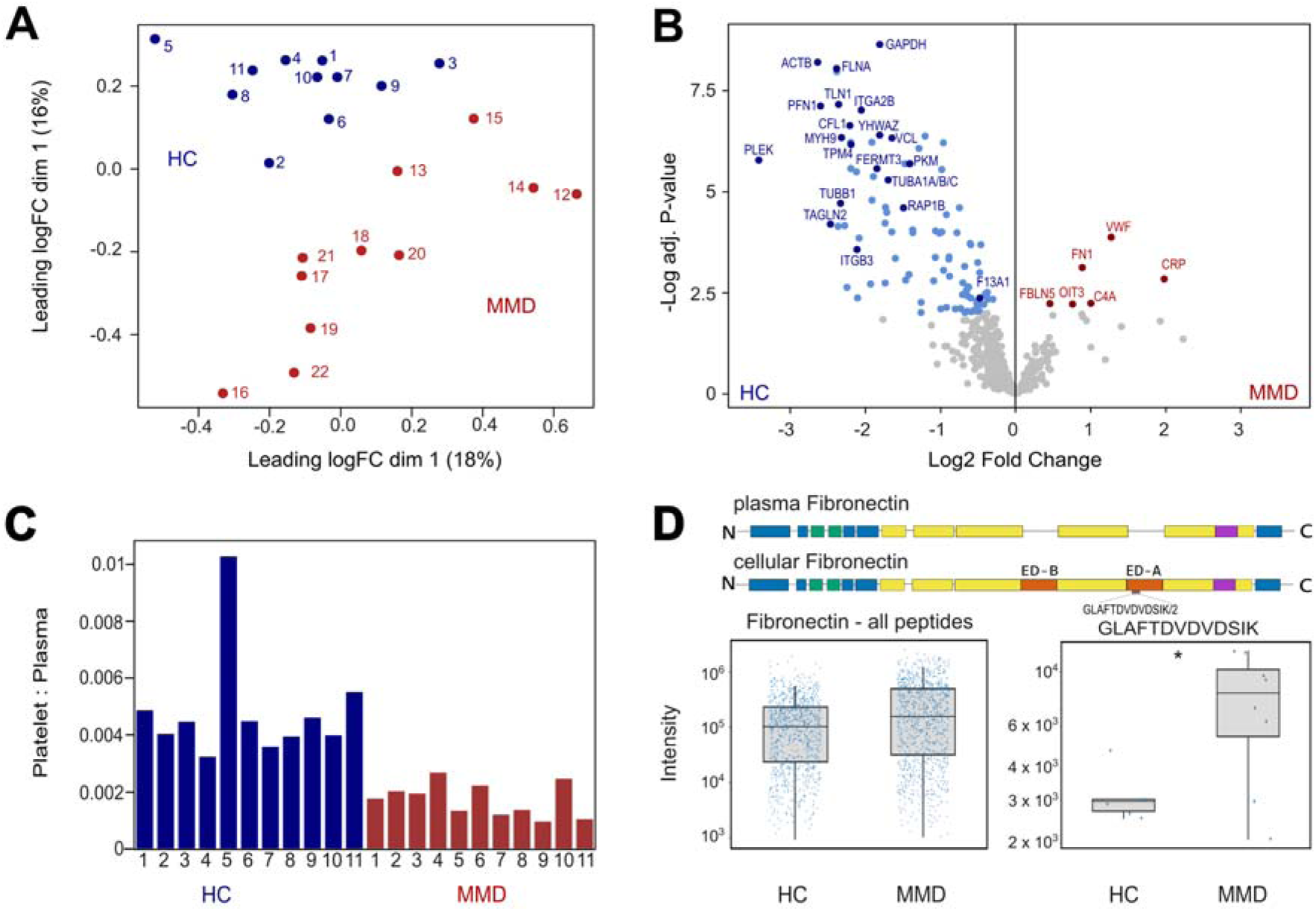
Plasma proteomics profiling reveals cFN1 as potential moyamoya disease biomarker. **(A)** Principal component analysis of healthy control (blue, HC) and moyamoya disease (red, MMD) plasma proteomics samples. (**B**) Volcano plot showing the differential protein abundance of MMD samples and healthy control plasma proteomics samples. Significant upregulated and downregulated proteins were indicated in red and blue, respectively. Platelet quality marker proteins defined by Geyer *et al.*^28^ are indicated in dark blue. (**C**) Platelet contaminant index for each individual sample according the method described by Geyer *et al.*^28^. (**D**) (*Top*) Schematic representation of the domain structure of plasma and cellular fibronectin. Regions with FN module type I, II, and III were indicated by blue, green and yellow rectangles, respectively, while the variable region in purple. (*Bottom*) Box plots showing the log2 peptide intensity in MMD patients and healthy controls (HC) of all detected FN1 peptides (*Left*) and the GLAFTDVDVDSIK peptide specific to the FN1-EDA domain (*Right*).

### Spatial single cell proteomics analysis of an occluded moyamoya artery

To gain more insight into the disease mechanisms underlying MMD we took advantage of unique brain autopsy material that was collected from patient 5. We used the MACSima™ imaging system to image a cross section of an occluded MMD artery at single cell resolution (**Fig. 2A**). This targeted proteomics approach allowed to stain with a commercial panel of 146 antibodies validated for FFPE tissue (see Methods), including several tumor and immune markers that fit well with MMD as an immune-related proliferative disease.^4^ In addition, we added six custom antibodies based on previous MMD research (ISG15, RNF213, IFN-γ, ACTA2, MMP2)^1,4,9,41^ and our plasma proteomics findings (FN1), generating unprecedented spatial protein expression data of the thickened intima and surrounding tissue layers (**Fig. 2B**). After cell segmentation and data quality filtering in a region of interest spanning the entire vessel, we obtained expression levels of 138 proteins across 2,362 cells.

**Figure 2.**
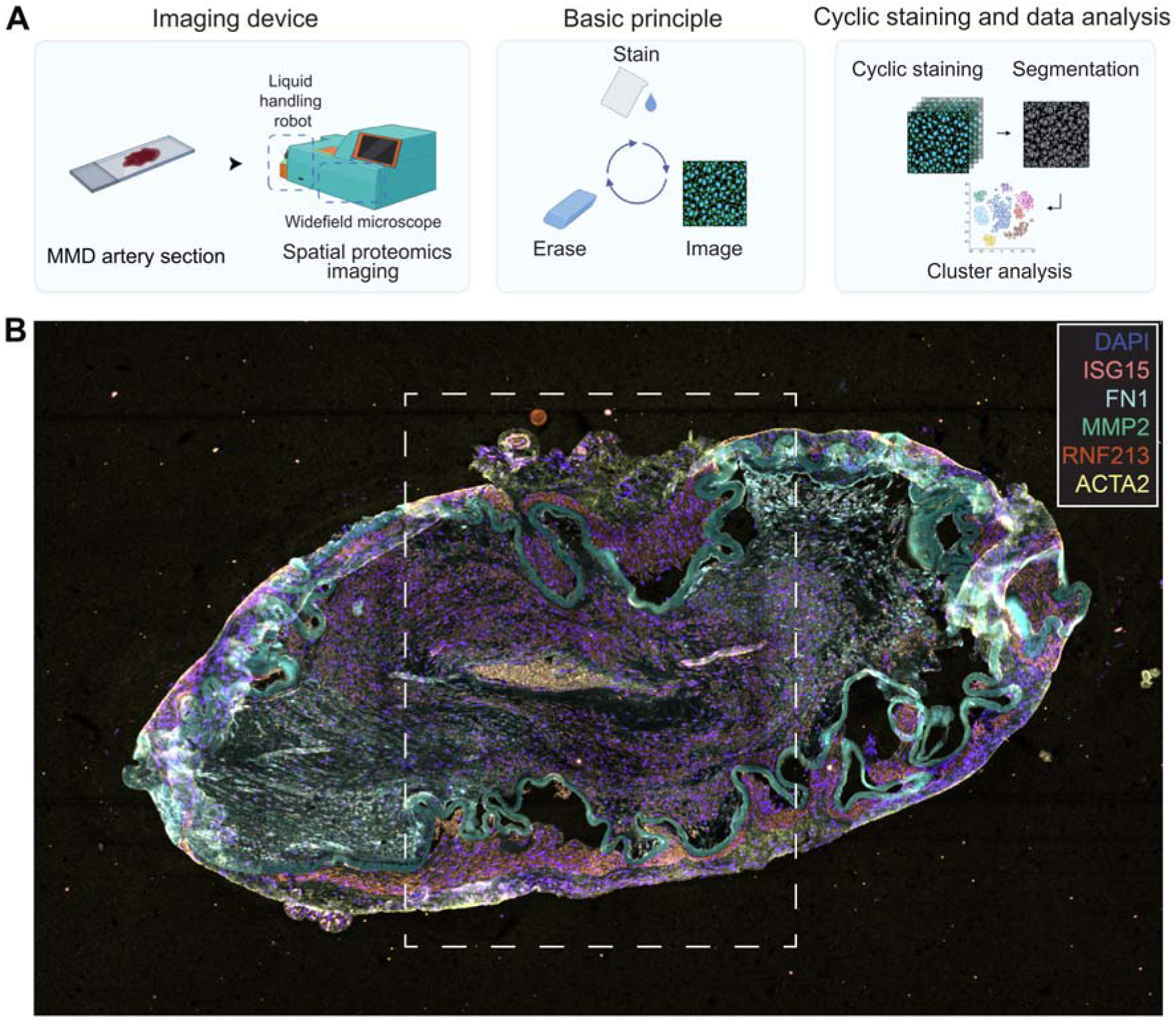
Spatial single cell proteomics analysis of an occluded moyamoya disease artery. **(A)** The MACSima™ Imaging System consists of a liquid handling robot and widefield microscope and operates by iterative immunofluorescent staining, sample washing, multi-field imaging, and signal erasure, using three fluorochrome-conjugated antibodies per cycle. Standard and novel image processing algorithms were employed to remove imaging artifacts and to maximize the signal-to-noise ratio. Further in-depth analysis allowed to segment the tissue images into single cells and cluster cells according to their expression profiles. Created with BioRender.com. **(B)** Representative image showing staining for DAPI (purple) and five different antibodies recognizing RNF213 (red), ACTA2 (yellow), MMP2 (green), FN1 (light blue), ISG15 (pink).

Single cell analysis using Seurat^23^ revealed five cell clusters with similar protein expression profiles. Mapping these cells back onto the sample image enabled us to explore the spatial distribution of these cell populations and allowed us to differentiate between five broad cell types of the vessel: a collagen rich outer layer consisting of connective tissue, collagen fibers and elastic fibers (adventitia), a middle layer made up of smooth muscle cells and elastic fibers (media) and an innermost layer which in a healthy artery consists of a thin layer of connective tissue (intima) and endothelial cells (endothelium) outlining the lumen (lumen) (**Fig. 3A**). In the MMD vessel, extreme thickening of the intima is clearly visible, almost completely obliterating the lumen. Nevertheless, the vessel appears to have been partially functional, as evidenced by red blood cells present in the residual lumen.

**Figure 3.**
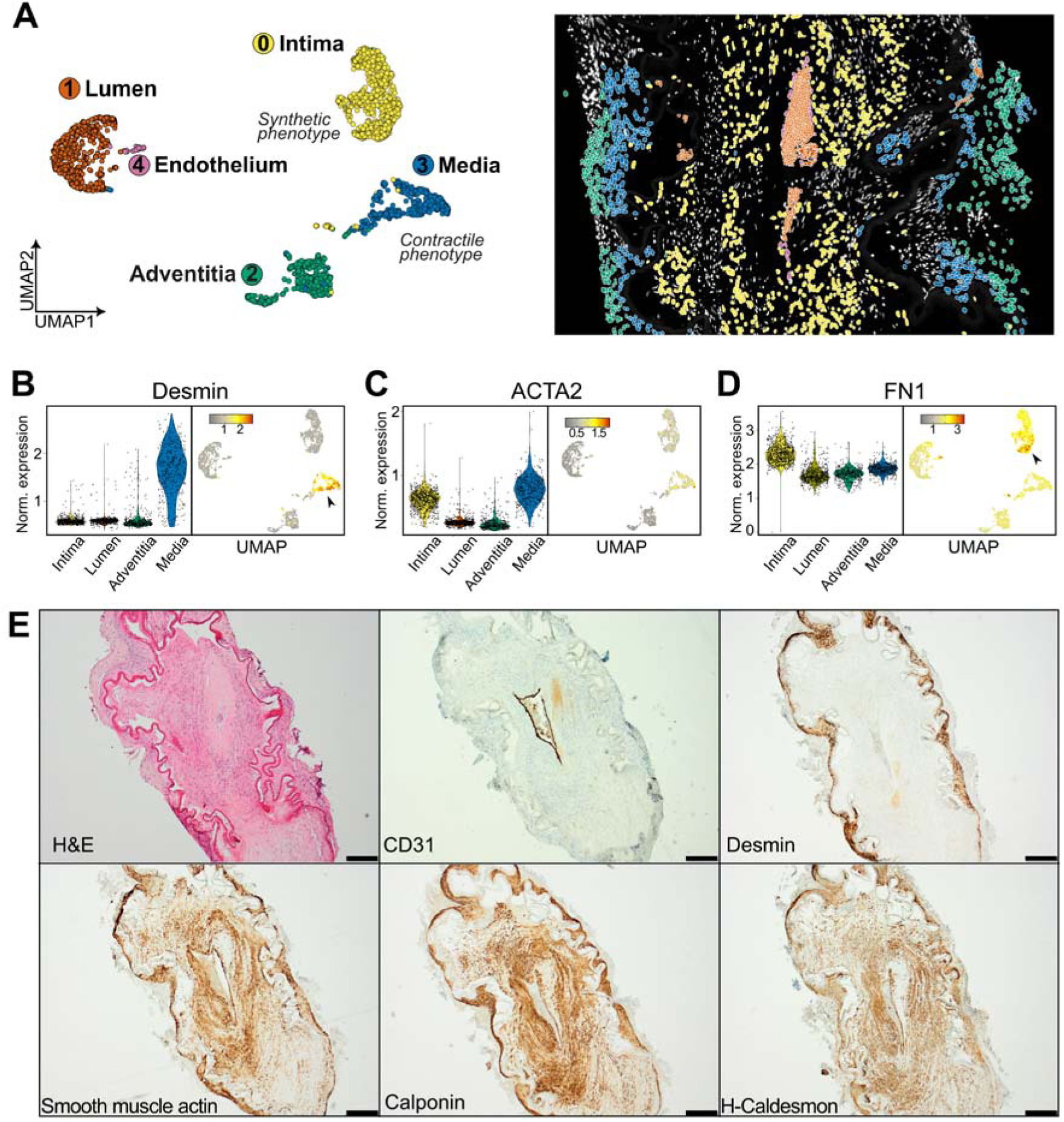
Phenotypic switching of intimal VSMCs. **(A)** Uniform Manifold Approximation and Projection (UMAP) analysis using Seurat^23^ for 2,362 high-quality cells with 138 features (antibody channels) revealed five cell clusters with similar protein expression profiles corresponding to the different tissue layers of the obliterated artery: (0) intima, (1) lumen, (2) adventitia, (3) media and (4) endothelium. **(B-D)** Normalized protein expression of DES (Desmin), ACTA2 (smooth muscle actin) and FN1 (Fibronectin1) shown in violin plots per cluster and per cell. **(E)** Hematoxylin & eosin (H&E) and immunohistochemistry validation staining for CD31, Desmin, smooth muscle actin, Calponin and H-Caldesmon.

The observed thickening of the intima is a key pathological hallmark of MMD resulting from abnormal infiltration and proliferation of vascular smooth muscle cells (VSMCs) in the intima.^16^ When comparing cell expression profiles in the intima and the media, we observed that intima VSMCs were relatively depleted in smooth muscle markers such as ACTA2 (smooth muscle actin) and especially DES (Desmin), while markers such as FN1 (fibronectin), KRT19 (cytokeratin 19), and GFAP (glial fibrillary acidic protein) showed high expression (**Fig. 3B-D** and **Supplementary Fig. 2**-5). This change in protein expression is in line with VSMC switching from a contractile to synthetic phenotype in the media and intima, respectively, as reported before.^1^ Validation by immunohistochemistry (IHC) confirmed the strong downregulation of DES expression in the intima, while cells in both layers stained positive for smooth muscle markers ACTA2, CNN1 (calponin-1) and CALD1 (caldesmon) (**Fig. 3E, Supplementary Fig. 6**).

Although MMD is not considered an inflammatory disease, a previous study reported infiltration of macrophages and T lymphocytes in the affected vessel wall.^1^ In line with immune triggers as potential second hit for MMD, we also assessed the presence of immune cells in our dataset using various immune cell markers present in our antibody panel. In total, we distinguished 223 immune cells that mainly resided in the intima, adventitia and lumen, and that clustered into four groups based on their protein expression (**Fig. 4A**). The majority of these immune cells (161 out of 223) resided in the adventitia and divided across two clusters. The adventitia-II cluster showed relatively high expression of T lymphocyte-associated markers CD8a, CD43^36^, CD45RO and CD45RB (**Fig. 4B, Supplementary Fig. 7**), while cells in the adventitia-I cluster were characterized by expression of CD204, a myeloid cell marker often associated with tissue-resident M2 macrophages (**Fig. 4C**).^42^ In contrast, immune cells infiltrating the intima showed elevated expression of HLA-DR-C and CD162 (**Fig. 4D**), markers for activated antigen presenting cells (APCs), including pro-inflammatory M1 macrophages.^43^ The third cluster comprised circulating immune cells detected in the residual lumen of the vessel that were characterized by expression of the granulocyte markers CD66 and CD15 (**Fig. 4E**). IHC using additional lymphoid cell markers (CD45, CD3, CD4, CD8 and CD20) confirmed the presence of lymphoid cell populations, such as T- and B-lymphocytes, in the outer adventitia layer (**Fig. 4F, Supplementary Fig. 6**), while CD68- positive macrophages were found both in the adventitia as well as in the intima where they were scattered between the VSMCs (**Fig. 4F**). Together, these data point to infiltration of macrophages, likely in a pro-inflammatory state, in the proliferating intima, while lymphoid cells remain in the adventitia.

**Figure 4.**
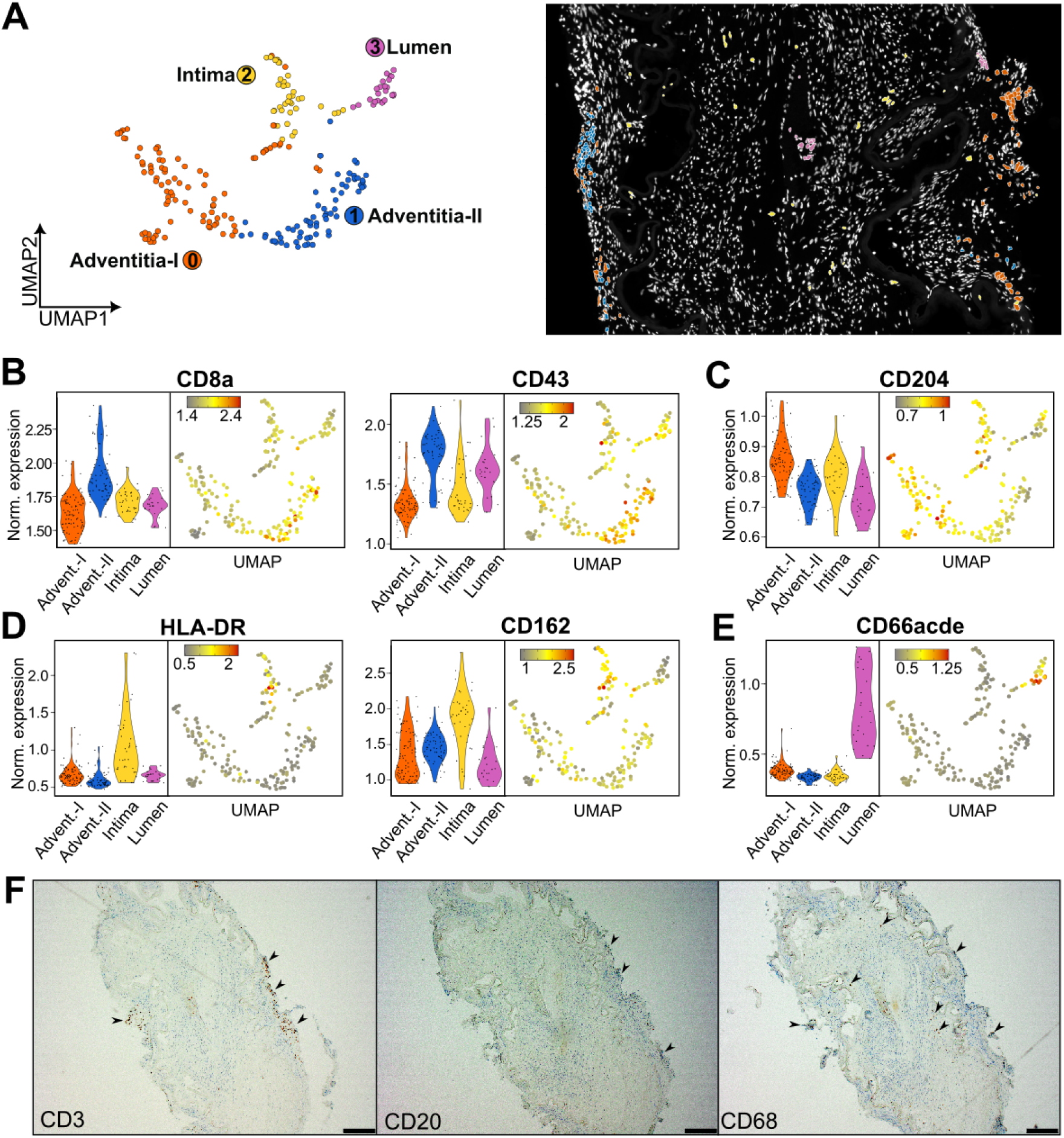
Immune cell infiltration in the occluded artery. **(A)** Uniform Manifold Approximation and Projection (UMAP) analysis using Seurat^23^ on a subset of 223 segmented immune cells revealed four clusters of cells present in the adventitia (cluster 0 and 1 or adventitia-I and -II, respectively), intima (cluster 2) and the lumen (cluster 3). **(B-E)** Normalized protein expression of CD8a, CD43, CD204, HLA-DR and CD162 and CD66acde shown in violin plots per cluster and per cell. (**F**) Immunohistochemistry validation staining for CD3, CD20 and CD68.

### Laser capture microdissection proteomics of a moyamoya disease artery

In addition to targeted imaging-based spatial proteomics, we also employed untargeted mass spectrometry (MS)-based proteomics to compare protein expression levels in the intima and media layer of the occluded MMD artery. To this end, we used laser capture microdissection to cut out pieces of tissue from the intima and media, followed by low-input quantitative LC-MS/MS analysis (**Fig. 5A-B**). Dimensionality reduction clearly separated intima and media samples (**Fig. 5C**) and differential analysis revealed four proteins (DES, DCN, HSPB6, SELENBP1) that were significantly downregulated in the intima compared to the media, while five proteins (TAGLN2, TTR, TSP1, FN1, POSTN) were significantly upregulated (**Fig. 5D**). Downregulation of DES corroborated the spatial proteomics and IHC results and together with HSPB6 indicates the loss of smooth muscle cell-specific protein expression.^1,44^ Along with upregulation of TAGLN2 (transgelin-2), an early smooth muscle marker, and upregulation of the ECM proteins such as FN1, POSTN (periostin) and TSP1 (thrombospondin-1), this change in protein expression pattern indicates a dedifferentiation process, in line with phenotypic switching from a contractile to a proliferative and synthetic phenotype of the VSMCs in the intima. Interestingly, the cFN1-EDA specific peptide GLAFTDVDVDSIK that was found upregulated in MMD patient plasma (**Fig. 1D**), was also higher abundant in the intimal layer together with another EDA-specific peptide (**Supplementary Fig. 8**). Together, these observations suggest a model where cFN1 expressed by proliferating intimal cells leak into the circulation near occlusive lesions, providing a rationale for plasma cFN1 as a moyamoya biomarker.

**Figure 5.**
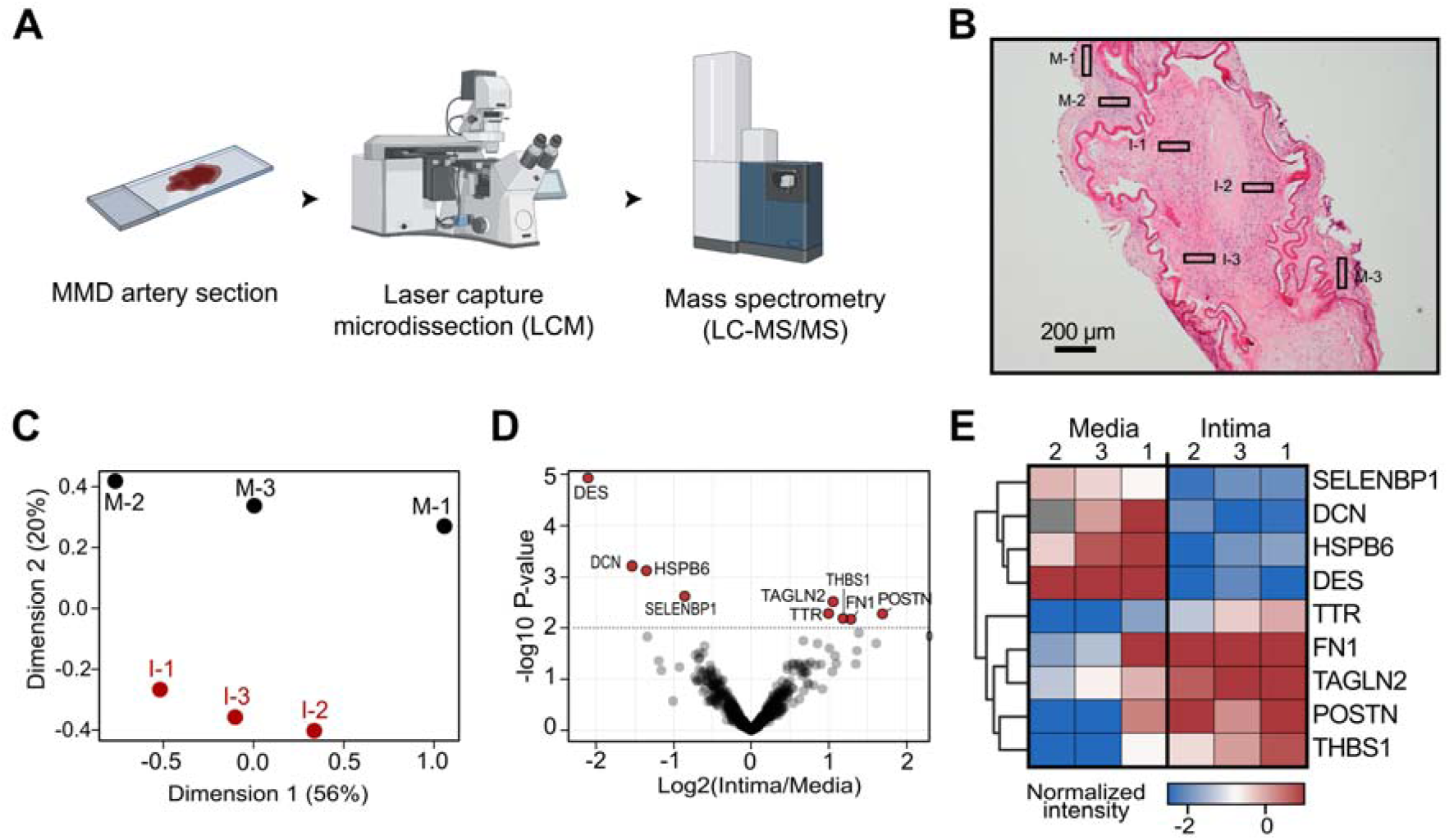
MS-based proteomics confirmed phenotypic switching of intimal VSMCs. (**A**) A fixed and paraffin-embedded sample of an occluded MMD brain artery was mounted on a glass slide for microscopy. Regions inside the intima (I-1, I-2 and I-3) and media (M-1, M-2 and M-3) of the obliterated vessel were selected and cut out by laser capture microdissection (LCM). Samples were then analyzed by liquid chromatography-tandem mass spectrometry (LC-MS/MS). (**B**) Hematoxylin and eosin (H&E) image indicating the regions selected for LCM and LC-MS/MS analysis. (**C**) Multidimensional scaling plot of all quantified proteins with percentages of explained data variance shown on the x and y axis. (**D**) Volcano plot showing nine proteins with significantly different expression levels between intima and media regions (*P* ≤ 0.01 and absolute log_2_ fold change > 1). (**E**) Heat map visualizing normalized intensities of the nine significant proteins in each sample after hierarchical clustering. A single missing intensity value is indicated in grey.

## Discussion

MMD is a chronic cerebrovascular occlusive disease, accompanied by an abnormal formation of collateral vessels of the vasculature at the base of brain.^16^ The complex pathogenesis calls for a multiomics approach to elucidate the yet unknown etiology of MMD, integrating genetic information on MMD susceptibility with mRNA- or protein-level information on potential dysregulated pathways and molecular triggers of the disease, in line with a previously suggested two-hit hypothesis.^4,45–47^ Multiomics research on MMD is however limited, so far focusing on analyses of plasma or CSF of MMD patients.^48–50^ Here, we report the genetic results of a moyamoya patient cohort in combination with plasma proteomics and a spatial proteomics analysis of an occluded artery of one of the patients.

In contrast to East-Asian cohorts in which the occurrence of MMD is largely driven by the *RNF213* variant p.(R4810K), our cohort depicted a more heterogeneous genetic architecture. Three patients carried a single nucleotide variant in RNF213, a known MMD susceptibility gene.^6,7,45^ *RNF213* is involved in multiple functions, among which a pivotal antimicrobial role was recently discovered.^8–10^ One of our patients, was a carrier of the founder variant p.(R4810K) of *RNF213*. This variant is reported to increase MMD risk by over 100-fold (reviewed in Mertens *et al.*^11^) although disease penetrance is still very low with only 0.5% of carriers of heterozygous p.(R4810K) developing MMD.^11^ Two other patients in our cohort carried a novel rare variant p.(A3927V) in the *RNF213* gene. A different variant affecting the same codon but leading to a different amino acid substitution p.(A3927T) was identified before in a patient from Caucasian descent.^16^ Similar to p.(R4810K) and many other MMD variants,^4^ p.(A3927V) is located in the E3 module of RNF213. Therefore, this new variant is potentially the genetic hit in these patients although further segregation and functional studies would be needed to establish the causal nature of this variant for MMD development.

Two patients with moyamoya-like angiopathy who also showed aorta dilatation, patent ductus arteriosus and congenital bilateral mydriasis (**Supplementary Material**) each carried a *de novo* pathogenic variant in *ACTA2*, the gene encoding alpha-smooth muscle actin.^51^ Pathogenic *ACTA2* variants have been reported to cause smooth muscle cell dysfunction and vascular disorders, including thoracic aortic aneurysms, aortic dissections, coronary heart disease, stroke and MMS.^17,51–54^ Interestingly, the specific p.(R179S) and p.(R179H) variants found here have been described in multiple Caucasian MMD patients.^17,51,54^

One patient in the cohort has been reported previously^18^ and was compound heterozygous for two variants in *SAMHD1*, p.(R226C) and a splice donor site mutation (c.1608+2 T>C). SAMHD1 functions as a deoxynucleoside triphosphate (dNTP) triphosphohydrolase that breaks down dNTPs, a mechanism that can restrict viral replication of HIV in certain immune cells.^55,56^ Pathogenic variants in *SAMHD1* can lead to Aicardi-Goutières syndrome, a rare neuroinflammatory disorder and type I interferonopathy,^18^ however, such *SAMHD1* mutations can also cause MMD, particularly in patients with bi-allelic variants.^18^ These impair the normal function of SAMHD1, disrupting regulation of dNTP levels and triggering a chronic type I interferon (IFN) response via DNA-sensing pathways like cGAS-STING.^56^ This IFN-driven inflammation damages endothelial cells leading to vasculopathy,^18,55–57^ likely promoting the development of conditions like MMA. Infants carrying pathogenic *SAMHD1* variants presenting with both Aicardi-Goutières syndrome and MMS have been described,^18,55,57^ however, the *SAMHD1* patient in our cohort does not exhibit Aicardi- Goutières syndrome, although he was found to have an elevated IFN-signaling gene expression.^18^

Next to pathogenic variants in single genes, MMD is frequently associated with other syndromes or systemic disorders such as Down syndrome (trisomy 21), sickle cell anemia, neurofibromatosis type-1, and other conditions.^58–60^ Our cohort includes one MMA patient with trisomy 21. In addition, patient 3 carried a small intragenic deletion in *NFIA*, a transcription factor gene*. NFIA* haploinsufficiency leads to *NFIA*-related disorder, an autosomal dominant neurodevelopmental condition characterized by a variable degree of cognitive impairment and non-specific dysmorphic features.^61^ Interestingly, MMA has been described in two patients with 1p32p31 chromosome deletion syndrome encompassing *NFIA*,^62,63^ suggesting that loss of one copy of *NFIA* can predispose individuals to MMA. Together, whole exome sequencing allowed to identify genetic susceptibility factors for most patients, illustrating the broad range of possible genetic triggers in MMA/MMD development.

In plasma samples from eleven patients, six proteins were found significantly upregulated compared to plasma of sex- and age-matched controls. Two of these proteins, CRP and C4A, are known to be elevated during an inflammatory state, with CRP as a widely used biomarker for detection and follow-up of systemic inflammation^64^ and C4A as part of the complement activation in the innate immune response.^65^ These findings are in line with previous observations and a state of low-grade inflammation that is emerging from recent studies,^36–39^ supporting the idea of an important immunological component in the development of MMD.^4^ Another group of upregulated proteins, OIT3, FBLN5 and FN1, are linked to cancer biology and uncontrolled cell growth.^33–35^ This fits with the proliferation of VSMCs and deposition of ECM in affected moyamoya arteries, especially since our spatial analysis also demonstrated upregulation of FN1 in the intimal layer of an occluded moyamoya artery. FN1 has two major isoforms, cFN1 and pFN1, which can be differentiated by the EDA and EDB domains that are specific for cFN1.^33^ Under normal conditions, cFN1 is locally excreted into surrounding tissues with no or minimal entry into the bloodstream, while pFN1 is the predominant isoform in plasma.^66^ Interestingly, detection of an EDA-specific peptide in moyamoya plasma suggests that the overall increase in circulating FN1 results at least partially from tissue leakage of cFN1. While cFN1 and its EDA/EDB domains have been explored as biomarkers for pneumological, vascular and especially oncological diseases,^33,66–70^ we here propose to extend their use as potential biomarker for MMD. A biomarker for MMD could facilitate early diagnosis of patients, especially in populations at risk, and help with prognosis and treatment monitoring.^71–73^ Among other proteins, previous studies proposed matrix metalloproteases (MMPs) and vascular endothelial growth factor (VEGF) as biomarkers upregulated in CSF, plasma and urine of moyamoya patients. Upregulation of MMPs and VEGF likely results from chronic ischemia in the brain and successful revascularization leads to a concomitant reduction in the levels of these markers. Interestingly, we found that upregulation of cFN1 in moyamoya plasma correlates with an upregulation of FN1 in the intimal layer of an occluded artery, suggesting that cFN1 might leak into the bloodstream near occlusive lesions. In this way, rather than informing on the ischemic state, cFN1 might act as a marker of artery tissue proliferation and correlate with the level of stenosis. This needs to be confirmed in further studies along with a validation cFN1 as a biomarker on larger patient cohorts using orthogonal methods for protein read-out.

In autopsy-derived material of patient 5 we used a targeted antibody-based spatial proteomics imaging approach and observed extensive infiltration and proliferation of cells in the intima layer of the moyamoya artery. In line with their reported origin as vascular smooth muscle cells,^1,74^ these cells stained positive for the smooth muscle marker ACTA2, Calponin and H-caldesmon, but showed a strong downregulation of desmin expression, which was also confirmed by IHC. Change in the expression of these marker proteins was reported before, as the result of phenotypic switching of these intima VSMCs from a contractile to a synthetic phenotype.^1^ Phenotypic switching of cells is a key mechanism in many vascular diseases.^75^ It describes the ability of cells, particularly VSMCs, to shift between different functional states in response to environmental stimuli or stress in the vascular environment, including vessel injury. This adaptability allows VSMCs to transition from a contractile phenotype which maintains vessel tone, to a synthetic phenotype promoting cellular growth, migration, and matrix production.^75^

A signature of increased ECM production was also apparent from an untargeted MS-based proteomics analysis, where we detected significant upregulation of ECM proteins FN1, TSP1 and POSTN in the intima compared to medial tissue. Interestingly, expression of FN1 by ACTA2-positive fibroblast-like VSMCs, first discovered in human and mouse atherosclerotic lesions,^76^ provides evidence for phenotypic switching of ingrowing VSMCs to this specific subtype of fibroblast-like VSMCs. Similarly, TSP-1 and POSTN promote intimal hyperplasia and ECM deposition characteristic for occluded moyamoya arteries.^77–80^ Next to ECM proteins, TAGLN2 (Transgelin-2) was found upregulated in intimal cells. TAGLN2 is an actin-binding protein constitutively expressed in immune cells, and is responsive to inflammatory signals such as LPS in APCs, indicating a role in activated macrophages and immune protection.^81,82^ TAGLN2 upregulation potentially reflects the inflammatory environment in MMD and hints again at dedifferentiation of intimal VSMCs.

The single cell resolution of our spatial data also allowed to detect immune cells infiltrating the occluded artery. In contrast to a previous study that reported the presence of both macrophages and T cells in the intima,^1^ we only observed macrophages in this layer, while T and B cells were detected in the adventitia, where residing lymphocytes are expected.^83^ This differential recruitment of immune cells warrants further investigation, especially in light of the emerging role of immune and infectious stimuli as potential triggers for MMD.^4^ Unlike atherosclerosis where both macrophages and T cells are commonly found in atherosclerotic plaques,^84–86^ moyamoya is not considered an inflammatory disease. However, the intimal infiltration of CD68-positive macrophages observed here by IHC suggests that low level inflammation cannot be excluded, especially since single cell imaging revealed a similar pattern of activated APCs in the intima. Due to lack of a CD68 antibody in the used MACSima panel we cannot exclude infiltration of other APCs next to macrophages, but in either case the pro-inflammatory (HLA-DR positive) state of these cells is in line with heightened inflammation markers recently found in peripheral blood of MMD patients.^38,39^ While the precise origin and function of these infiltrating APCs remains unclear, they could disturb the normal crosstalk between endothelial cells and VSMCs for instance via nitric oxide,^46^ thereby promoting VSMC phenotypic switching. Of note, in addition to being a marker of phenotypic switching of VSMCs, FN1, a protein that is upregulated in the intimal layer of the artery in our data, can be upregulated in an inflammatory environment. FN1 can interact with immune cells, including macrophages and fibroblasts, to promote the recruitment of inflammatory cells to the vessel wall,^87^ further supporting its potential as a disease-related biomarker. Together, the spatial analysis described here provides an unprecedented view on an occluded MMD artery at the single cell level, unveiling VSMC phenotypic switching and inflammatory immune cell infiltration in the intima layer, providing a rationale for cellular fibronectin as potential biomarker for MMD in plasma.

## Acknowledgements

The authors would like to thank the patients and families who participated in this study. We thank Martin Guilliams, Charlotte Scott and the VIB Spatial Catalyst for advice and access to the MACSima™ Imaging System. We acknowledge help from Janick Mathys on the single cell data analysis and we thank the VIB Proteomics Core and Lode Denolf for help with analysis of the mass spectrometry proteomics samples.

## Author contributions

Investigation: CA, JVD; Formal analysis and data curation: JuM and PW; Visualization: CA, JuM and PW Resources: FV, WDS, EVH, DH; Writing – Original Draft: CA, JoM, PW, BD and FI; Writing – Review & Editing: JuM, FV, ED, AS, SL, WDS, EF, KG, EVH, JVD and DH; Conceptualization: DH, EVH, BD and FI; Supervision: BD and FI. All authors read and approved the final manuscript.

## Data availability

The mass spectrometry proteomics data of the laser capture microdissection (LCM) and plasma proteomics have been deposited to the ProteomeXchange Consortium (http://proteomecentral.proteomexchange.org) via the PRIDE partner repository^24^ with the dataset identifiers PXD057183 and PXD063236, respectively. All code and data tables used to analyze the spatial proteomics data (image, single cell, differential protein statistics) is available in a GitHub repository archived on Zenodo https://doi.org/10.5281/zenodo.17158033. An overview of patient medical histories is available from the corresponding author upon reasonable request.

## Ethics declarations

### Ethics approval and consent to participate

The study was approved by the Ethics Committee of UZ Gent (EC: 2019/1430). All procedures were in accordance with the ethical standards of the Ethics Committee of UZ Gent and with the 1964 Helsinki declaration and its later amendments or comparable ethical standards. Written informed consents for multiomics analysis and publication of results were obtained from all patients.

## Supplementary Information

Contents:

- Supplementary Figures 1-8
- Supplementary Table 1
- Supplementary Methods

## References

1. Masuda J, Ogata J, Yutani C. Smooth muscle cell proliferation and localization of macrophages and T cells in the occlusive intracranial major arteries in moyamoya disease. Stroke. Dec 1993;24(12):1960–7. doi:10.1161/01.str.24.12.1960

2. Kuroda S, Fujimura M, Takahashi J, et al. Diagnostic Criteria for Moyamoya Disease - 2021 Revised Version. Neurol Med Chir (Tokyo). Jul 15 2022;62(7):307–312. doi:10.2176/jns-nmc.2022-0072

3. Scott RM, Smith ER. Moyamoya disease and moyamoya syndrome. N Engl J Med. Mar 19 2009;360(12):1226–37. doi:10.1056/NEJMra0804622

4. Asselman C, Hemelsoet D, Eggermont D, Dermaut B, Impens F. Moyamoya disease emerging as an immune-related angiopathy. Trends Mol Med. Nov 2022;28(11):939–950. doi:10.1016/j.molmed.2022.08.009

5. Kuriyama S, Kusaka Y, Fujimura M, et al. Prevalence and clinicoepidemiological features of moyamoya disease in Japan: findings from a nationwide epidemiological survey. Stroke. Jan 2008;39(1):42–7. doi:10.1161/STROKEAHA.107.490714

6. Kamada F, Aoki Y, Narisawa A, et al. A genome-wide association study identifies RNF213 as the first Moyamoya disease gene. J Hum Genet. Jan 2011;56(1):34–40. doi:10.1038/jhg.2010.132

7. Liu W, Morito D, Takashima S, et al. Identification of RNF213 as a susceptibility gene for moyamoya disease and its possible role in vascular development. PLoS One. 2011;6(7):e22542. doi:10.1371/journal.pone.0022542

8. Otten EG, Werner E, Crespillo-Casado A, et al. Ubiquitylation of lipopolysaccharide by RNF213 during bacterial infection. Nature. Jun 2021;594(7861):111-116. doi:10.1038/s41586-021-03566-4

9. Thery F, Martina L, Asselman C, et al. Ring finger protein 213 assembles into a sensor for ISGylated proteins with antimicrobial activity. Nat Commun. Oct 1 2021;12(1):5772. doi:10.1038/s41467-021-26061-w

10. Houzelstein D, Simon-Chazottes D, Batista L, et al. The ring finger protein 213 gene (Rnf213) contributes to Rift Valley fever resistance in mice. Mamm Genome. Feb 2021;32(1):30–37. doi:10.1007/s00335-020-09856-y

11. Mertens R, Graupera M, Gerhardt H, et al. The Genetic Basis of Moyamoya Disease. Transl Stroke Res. Feb 2022;13(1):25–45. doi:10.1007/s12975-021-00940-2

12. Strunk D, Diehl RR, Veltkamp R, Meuth SG, Kraemer M. Progression of initially unilateral Moyamoya angiopathy in Caucasian Europeans. J Neurol. Sep 2023;270(9):4415–4422. doi:10.1007/s00415-023-11793-0

13. Church EW, Bell-Stephens TE, Bigder MG, Gummidipundi S, Han SS, Steinberg GK. Clinical Course of Unilateral Moyamoya Disease. Neurosurgery. Nov 16 2020;87(6):1262–1268. doi:10.1093/neuros/nyaa284

14. Savolainen M, Mustanoja S, Pekkola J, et al. Moyamoya angiopathy: long-term follow-up study in a Finnish population. J Neurol. Mar 2019;266(3):574–581. doi:10.1007/s00415-018-9154-7

15. Ok T, Jung YH, Kim J, et al. RNF213 R4810K Variant in Suspected Unilateral Moyamoya Disease Predicts Contralateral Progression. J Am Heart Assoc. Aug 2 2022;11(15):e025676. doi:10.1161/JAHA.122.025676

16. 16. Research Committee on the P, Treatment of Spontaneous Occlusion of the Circle of W, Health Labour Sciences Research Grant for Research on Measures for Infractable D. Guidelines for diagnosis and treatment of moyamoya disease (spontaneous occlusion of the circle of Willis). Neurol Med Chir (Tokyo). 2012;52(5):245–66. doi:10.2176/nmc.52.245

17. Plagnol V, Curtis J, Epstein M, et al. A robust model for read count data in exome sequencing experiments and implications for copy number variant calling. Bioinformatics. Nov 1 2012;28(21):2747–54. doi:10.1093/bioinformatics/bts526

18. Richards S, Aziz N, Bale S, et al. Standards and guidelines for the interpretation of sequence variants: a joint consensus recommendation of the American College of Medical Genetics and Genomics and the Association for Molecular Pathology. Genet Med. May 2015;17(5):405–24. doi:10.1038/gim.2015.30

19. Schindelin J, Arganda-Carreras I, Frise E, et al. Fiji: an open-source platform for biological-image analysis. Nat Methods. Jun 28 2012;9(7):676–82. doi:10.1038/nmeth.2019

20. Bankhead P, Loughrey MB, Fernandez JA, et al. QuPath: Open source software for digital pathology image analysis. Sci Rep. Dec 4 2017;7(1):16878. doi:10.1038/s41598-017-17204-5

21. Stringer C, Wang T, Michaelos M, Pachitariu M. Cellpose: a generalist algorithm for cellular segmentation. Nat Methods. Jan 2021;18(1):100–106. doi:10.1038/s41592-020-01018-x

22. Berg S, Kutra D, Kroeger T, et al. ilastik: interactive machine learning for (bio)image analysis. Nat Methods. Dec 2019;16(12):1226–1232. doi:10.1038/s41592-019-0582-9

23. Satija R, Farrell JA, Gennert D, Schier AF, Regev A. Spatial reconstruction of single- cell gene expression data. Nat Biotechnol. May 2015;33(5):495–502. doi:10.1038/nbt.3192

24. Perez-Riverol Y, Bai J, Bandla C, et al. The PRIDE database resources in 2022: a hub for mass spectrometry-based proteomics evidences. Nucleic Acids Res. Jan 7 2022;50(D1):D543–D552. doi:10.1093/nar/gkab1038

25. Guey S, Kraemer M, Herve D, et al. Rare RNF213 variants in the C-terminal region encompassing the RING-finger domain are associated with moyamoya angiopathy in Caucasians. Eur J Hum Genet. Aug 2017;25(8):995–1003. doi:10.1038/ejhg.2017.92

26. Lauer A, Speroni SL, Patel JB, et al. Cerebrovascular Disease Progression in Patients With ACTA2 Arg179 Pathogenic Variants. Neurology. Jan 26 2021;96(4):e538–e552. doi:10.1212/WNL.0000000000011210

27. Karla AR, Pinard A, Boerio ML, et al. SAMHD1 compound heterozygous rare variants associated with moyamoya and mitral valve disease in the absence of other features of Aicardi-Goutieres syndrome. Am J Med Genet A. Apr 2024;194(4):e63486. doi:10.1002/ajmg.a.63486

28. Geyer PE, Voytik E, Treit PV, et al. Plasma Proteome Profiling to detect and avoid sample-related biases in biomarker studies. EMBO Mol Med. Nov 7 2019;11(11):e10427. doi:10.15252/emmm.201910427

29. Canavero I, Vetrano IG, Zedde M, et al. Clinical Management of Moyamoya Patients. J Clin Med. Aug 17 2021;10(16)doi:10.3390/jcm10163628

30. Atiq F, O’Donnell JS. Novel functions for von Willebrand factor. Blood. Sep 19 2024;144(12):1247–1256. doi:10.1182/blood.2023021915

31. Black S, Kushner I, Samols D. C-reactive Protein. J Biol Chem. Nov 19 2004;279(47):48487–90. doi:10.1074/jbc.R400025200

32. Simoni L, Presumey J, van der Poel CE, et al. Complement C4A Regulates Autoreactive B Cells in Murine Lupus. Cell Rep. Nov 3 2020;33(5):108330. doi:10.1016/j.celrep.2020.108330

33. Hall RC, Vaidya AM, Schiemann WP, Pan Q, Lu ZR. RNA-Seq Analysis of Extradomain A and Extradomain B Fibronectin as Extracellular Matrix Markers for Cancer. Cells. Feb 21 2023;12(5)doi:10.3390/cells12050685

34. Heo JH, Song JY, Jeong JY, et al. Fibulin-5 is a tumour suppressor inhibiting cell migration and invasion in ovarian cancer. J Clin Pathol. Feb 2016;69(2):109–16. doi:10.1136/jclinpath-2015-203129

35. Wen J, Yang S, Yan G, et al. Increased OIT3 in macrophages promotes PD-L1 expression and hepatocellular carcinogenesis via NF-kappaB signaling. Exp Cell Res. Jul 15 2023;428(2):113651. doi:10.1016/j.yexcr.2023.113651

36. Han W, Qiao Y, Zhang H, et al. Circulating sortilin levels are associated with inflammation in patients with moyamoya disease. Metab Brain Dis. Jan 2021;36(1):103–109. doi:10.1007/s11011-020-00616-0

37. Kim JH, Jeon H, Kim M, et al. Chemical and perfusion markers as predictors of moyamoya disease progression and complication types. Sci Rep. Jan 2 2024;14(1):56. doi:10.1038/s41598-023-47984-y

38. Liu C, Mou S, Zhang B, et al. Innate Immune Cell Profiling in Peripheral Blood Mononuclear Cells of Patients with Moyamoya Disease. Inflammation. Dec 13 2024;doi:10.1007/s10753-024-02201-4

39. Liu E, Liu C, Jin L, et al. Clinical value of the systemic immune-inflammation index in moyamoya disease. Front Neurol. 2023;14:1123951. doi:10.3389/fneur.2023.1123951

40. To WS, Midwood KS. Plasma and cellular fibronectin: distinct and independent functions during tissue repair. Fibrogenesis Tissue Repair. Sep 16 2011;4:21. doi:10.1186/1755-1536-4-21

41. Sesen J, Driscoll J, Moses-Gardner A, Orbach DB, Zurakowski D, Smith ER. Non- invasive Urinary Biomarkers in Moyamoya Disease. Front Neurol. 2021;12:661952. doi:10.3389/fneur.2021.661952

42. Uchida Y, Nishitai G, Kikuchi K, Shibuya T, Asano K, Tanaka M. CD204-positive monocytes and macrophages ameliorate septic shock by suppressing proinflammatory cytokine production in mice. Biochem Biophys Rep. Sep 2020;23:100791. doi:10.1016/j.bbrep.2020.100791

43. Strizova Z, Benesova I, Bartolini R, et al. M1/M2 macrophages and their overlaps - myth or reality? Clin Sci (Lond). Aug 14 2023;137(15):1067–1093. doi:10.1042/CS20220531

44. Dreiza CM, Komalavilas P, Furnish EJ, et al. The small heat shock protein, HSPB6, in muscle function and disease. Cell Stress Chaperones. Jan 2010;15(1):1–11. doi:10.1007/s12192-009-0127-8

45. Mineharu Y, Liu W, Inoue K, et al. Autosomal dominant moyamoya disease maps to chromosome 17q25.3. Neurology. Jun 10 2008;70(24 Pt 2):2357-63. doi:10.1212/01.wnl.0000291012.49986.f9

46. Mineharu Y, Miyamoto S. RNF213 and GUCY1A3 in Moyamoya Disease: Key Regulators of Metabolism, Inflammation, and Vascular Stability. Front Neurol. 2021;12:687088. doi:10.3389/fneur.2021.687088

47. Bang OY, Fujimura M, Kim SK. The Pathophysiology of Moyamoya Disease: An Update. J Stroke. Jan 2016;18(1):12–20. doi:10.5853/jos.2015.01760

48. Guo Q, Wang QN, Li J, et al. Proteomic and metabolomic characterizations of moyamoya disease patient sera. Brain Behav. Dec 2023;13(12):e3328. doi:10.1002/brb3.3328

49. Carrozzini T, Pollaci G, Gorla G, et al. Proteome Profiling of the Dura Mater in Patients with Moyamoya Angiopathy. Int J Mol Sci. Jul 7 2023;24(13)doi:10.3390/ijms241311194

50. Gatti L, Bersano A, Gorla G, Pollaci G, Carrozzini T, Potenza A. Multi-omic approaches for biomarker discovery in Moyamoya disease. Clinical and Translational Discovery. 2024/04/01 2024;4(2):e270. 10.1002/ctd2.270

51. Cuoco JA, Busch CM, Klein BJ, et al. ACTA2 Cerebral Arteriopathy: Not Just a Puff of Smoke. Cerebrovasc Dis. 2018;46(3-4):161–171. doi:10.1159/000493863

52. Guo DC, Papke CL, Tran-Fadulu V, et al. Mutations in smooth muscle alpha-actin (ACTA2) cause coronary artery disease, stroke, and Moyamoya disease, along with thoracic aortic disease. Am J Hum Genet. May 2009;84(5):617–27. doi:10.1016/j.ajhg.2009.04.007

53. Milewicz DM, Ostergaard JR, Ala-Kokko LM, et al. De novo ACTA2 mutation causes a novel syndrome of multisystemic smooth muscle dysfunction. Am J Med Genet A. Oct 2010;152A(10):2437-43. doi:10.1002/ajmg.a.33657

54. Roder C, Peters V, Kasuya H, et al. Analysis of ACTA2 in European Moyamoya disease patients. Eur J Paediatr Neurol. Mar 2011;15(2):117–22. doi:10.1016/j.ejpn.2010.09.002

55. Senju C, Nakazawa Y, Shimada M, et al. Aicardi-Goutieres syndrome with SAMHD1 deficiency can be diagnosed by unscheduled DNA synthesis test. Front Pediatr. 2022;10:1048002. doi:10.3389/fped.2022.1048002

56. Coggins SA, Mahboubi B, Schinazi RF, Kim B. SAMHD1 Functions and Human Diseases. Viruses. Mar 31 2020;12(4)doi:10.3390/v12040382

57. Xin B, Jones S, Puffenberger EG, et al. Homozygous mutation in SAMHD1 gene causes cerebral vasculopathy and early onset stroke. Proc Natl Acad Sci U S A. Mar 29 2011;108(13):5372–7. doi:10.1073/pnas.1014265108

58. Abdelgadir A, Akram H, Dick MH, et al. A Better Understanding of Moyamoya in Trisomy 21: A Systematic Review. Cureus. Mar 2022;14(3):e23502. doi:10.7759/cureus.23502

59. Vargiami E, Sapountzi E, Samakovitis D, et al. Moyamoya syndrome and neurofibromatosis type 1. Ital J Pediatr. Jun 21 2014;40:59. doi:10.1186/1824-7288-40-59

60. Phi JH, Wang KC, Lee JY, Kim SK. Moyamoya Syndrome: A Window of Moyamoya Disease. J Korean Neurosurg Soc. Jun 2015;57(6):408–14. doi:10.3340/jkns.2015.57.6.408

61. Dini G, Verrotti A, Gorello P, et al. NFIA haploinsufficiency: case series and literature review. Front Pediatr. 2023;11:1292654. doi:10.3389/fped.2023.1292654

62. Prontera P, Rogaia D, Mencarelli A, et al. Juvenile Moyamoya and Craniosynostosis in a Child with Deletion 1p32p31: Expanding the Clinical Spectrum of 1p32p31 Deletion Syndrome and a Review of the Literature. Int J Mol Sci. Sep 17 2017;18(9)doi:10.3390/ijms18091998

63. Pires de Oliveira-Sobrinho R, Bispo LM, Heleno JL, et al. Microdeletion 1p32p31 Presenting with Moyamoya Disease and Incomplete Hippocampal Inversion. Mol Syndromol. Jun 2024;15(3):225–231. doi:10.1159/000535240

64. Levinson T, Wasserman A. C-Reactive Protein Velocity (CRPv) as a New Biomarker for the Early Detection of Acute Infection/Inflammation. Int J Mol Sci. Jul 22 2022;23(15)doi:10.3390/ijms23158100

65. Heurich M, Focking M, Cotter D. Complement C4, C4A and C4a - What they do and how they differ. Brain Behav Immun Health. Aug 2024;39:100809. doi:10.1016/j.bbih.2024.100809

66. Lieverse RIY, Marcus D, van der Wiel AMA, et al. Human fibronectin extra domain B as a biomarker for targeted therapy in cancer. Mol Oncol. Jul 2020;14(7):1555–1568. doi:10.1002/1878-0261.12705

67. Castellanos M, Leira R, Serena J, et al. Plasma cellular-fibronectin concentration predicts hemorrhagic transformation after thrombolytic therapy in acute ischemic stroke. Stroke. Jul 2004;35(7):1671–6. doi:10.1161/01.STR.0000131656.47979.39

68. Lemanska-Perek A, Krzyzanowska-Golab D, Pupek M, Klimeczek P, Witkiewicz W, Katnik-Prastowska I. Analysis of Soluble Molecular Fibronectin-Fibrin Complexes and EDA-Fibronectin Concentration in Plasma of Patients with Atherosclerosis. Inflammation. Jun 2016;39(3):1059–68. doi:10.1007/s10753-016-0336-0

69. Hansen AH, Breisnes HW, Prior TS, et al. A serologically assessed neo-epitope biomarker of cellular fibronectin degradation is related to pulmonary fibrosis. Clin Biochem. Aug 2023;118:110599. doi:10.1016/j.clinbiochem.2023.110599

70. Li F, Hooper AT, Golas J, Chang CB, Neubert H, King L. Evaluation of EDB Fibronectin in Plasma, Patient-Derived Xenograft Formalin-Fixed Paraffin-Embedded and Fresh Frozen Tumor Tissues Using Immunoaffinity LC-MS/MS. J Proteome Res. Oct 7 2022;21(10):2331–2340. doi:10.1021/acs.jproteome.2c00182

71. Ferriero DM, Fullerton HJ, Bernard TJ, et al. Management of Stroke in Neonates and Children: A Scientific Statement From the American Heart Association/American Stroke Association. Stroke. Mar 2019;50(3):e51–e96. doi:10.1161/STR.0000000000000183

72. 72. Smith ER. Moyamoya Biomarkers. J Korean Neurosurg Soc. Jun 2015;57(6):415–21. doi:10.3340/jkns.2015.57.6.415

73. Lehman LL, Kaseka ML, Stout J, et al. Pediatric Moyamoya Biomarkers: Narrowing the Knowledge Gap. Semin Pediatr Neurol. Oct 2022;43:101002. doi:10.1016/j.spen.2022.101002

74. Milewicz DM, Kwartler CS, Papke CL, Regalado ES, Cao J, Reid AJ. Genetic variants promoting smooth muscle cell proliferation can result in diffuse and diverse vascular diseases: evidence for a hyperplastic vasculomyopathy. Genet Med. Apr 2010;12(4):196–203. doi:10.1097/GIM.0b013e3181cdd687

75. Chen R, McVey DG, Shen D, Huang X, Ye S. Phenotypic Switching of Vascular Smooth Muscle Cells in Atherosclerosis. J Am Heart Assoc. Oct 17 2023;12(20):e031121. doi:10.1161/JAHA.123.031121

76. Wirka RC, Wagh D, Paik DT, et al. Atheroprotective roles of smooth muscle cell phenotypic modulation and the TCF21 disease gene as revealed by single-cell analysis. Nat Med. Aug 2019;25(8):1280–1289. doi:10.1038/s41591-019-0512-5

77. Stein JJ, Iwuchukwu C, Maier KG, Gahtan V. Thrombospondin-1-induced vascular smooth muscle cell migration and proliferation are functionally dependent on microRNA-21. Surgery. Feb 2014;155(2):228–33. doi:10.1016/j.surg.2013.08.003

78. Moura R, Tjwa M, Vandervoort P, Cludts K, Hoylaerts MF. Thrombospondin-1 activates medial smooth muscle cells and triggers neointima formation upon mouse carotid artery ligation. Arterioscler Thromb Vasc Biol. Oct 2007;27(10):2163–9. doi:10.1161/ATVBAHA.107.151282

79. Alesutan I, Henze LA, Boehme B, et al. Periostin Augments Vascular Smooth Muscle Cell Calcification via beta-Catenin Signaling. Biomolecules. Aug 21 2022;12(8)doi:10.3390/biom12081157

80. Kim CW, Pokutta-Paskaleva A, Kumar S, et al. Disturbed Flow Promotes Arterial Stiffening Through Thrombospondin-1. Circulation. Sep 26 2017;136(13):1217–1232. doi:10.1161/CIRCULATIONAHA.116.026361

81. Kim HR, Park JS, Karabulut H, Yasmin F, Jun CD. Transgelin-2: A Double-Edged Sword in Immunity and Cancer Metastasis. Front Cell Dev Biol. 2021;9:606149. doi:10.3389/fcell.2021.606149

82. Na BR, Kim HR, Piragyte I, et al. TAGLN2 regulates T cell activation by stabilizing the actin cytoskeleton at the immunological synapse. J Cell Biol. Apr 13 2015;209(1):143–62. doi:10.1083/jcb.201407130

83. Cautivo KM, Steer CA, Molofsky AB. Immune outposts in the adventitia: One foot in sea and one on shore. Curr Opin Immunol. Jun 2020;64:34–41. doi:10.1016/j.coi.2020.03.005

84. Saigusa R, Winkels H, Ley K. T cell subsets and functions in atherosclerosis. Nat Rev Cardiol. Jul 2020;17(7):387–401. doi:10.1038/s41569-020-0352-5

85. Fox BM, Dorschel KB, Lawton MT, Wanebo JE. Pathophysiology of Vascular Stenosis and Remodeling in Moyamoya Disease. Front Neurol. 2021;12:661578. doi:10.3389/fneur.2021.661578

86. Tchernychev B, Nitschke Y, Chu D, et al. Inhibition of Vascular Smooth Muscle Cell Proliferation by ENPP1: The Role of CD73 and the Adenosine Signaling Axis. Cells. Jun 29 2024;13(13)doi:10.3390/cells13131128

87. Wang H, Zhang J, Li H, et al. FN1 is a prognostic biomarker and correlated with immune infiltrates in gastric cancers. Front Oncol. 2022;12:918719. doi:10.3389/fonc.2022.918719

