## Supplementary Material for "Increased plasma fibronectin mirrors intimal phenotypic switching of vascular smooth muscle cells in moyamoya arteriopathy"

#### Table of contents


| | |
|---|---|
| Supplementary Figures | Supplementary Figures 1-8. |
| Supplementary Table 1 | Antibodies used for immunofluorescence staining in MACSima™. |
| Supplementary Methods | Plasma and laser capture microdissection proteomics and proteomics data analysis. |

#### Supplementary figures

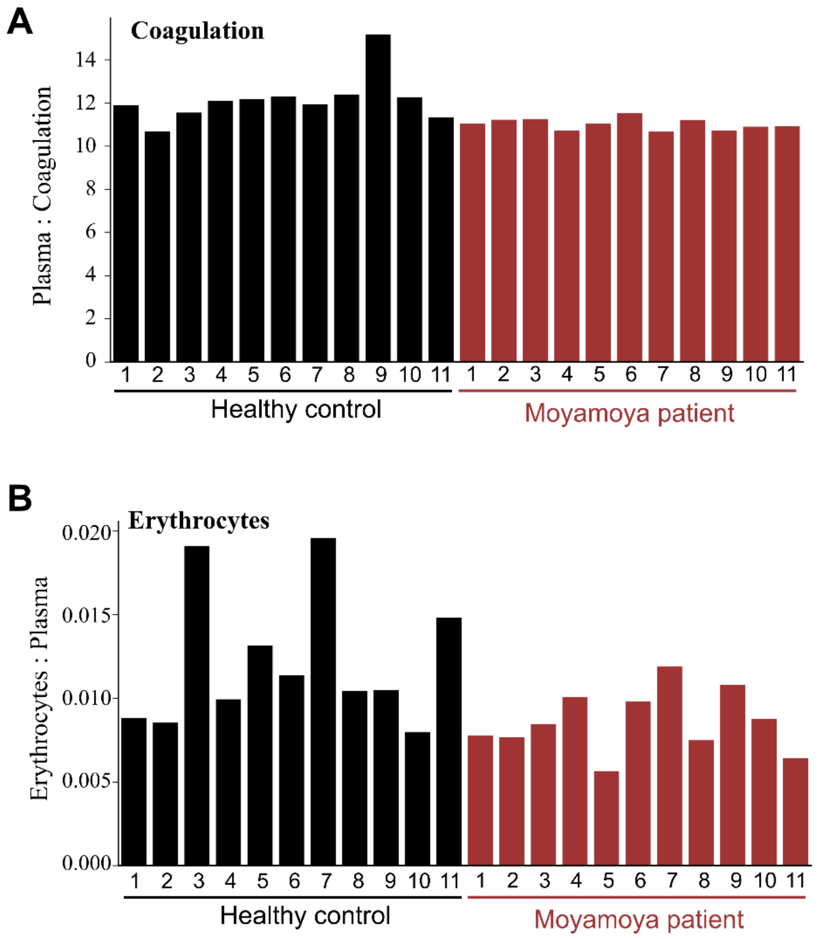


**Supplementary Figure 1.** Coagulation and erythrocyte cocontaminant index for each individual plasma sample calculated according to Geyer et al. (2019, *EMBO Mol Med* 11:e10427).

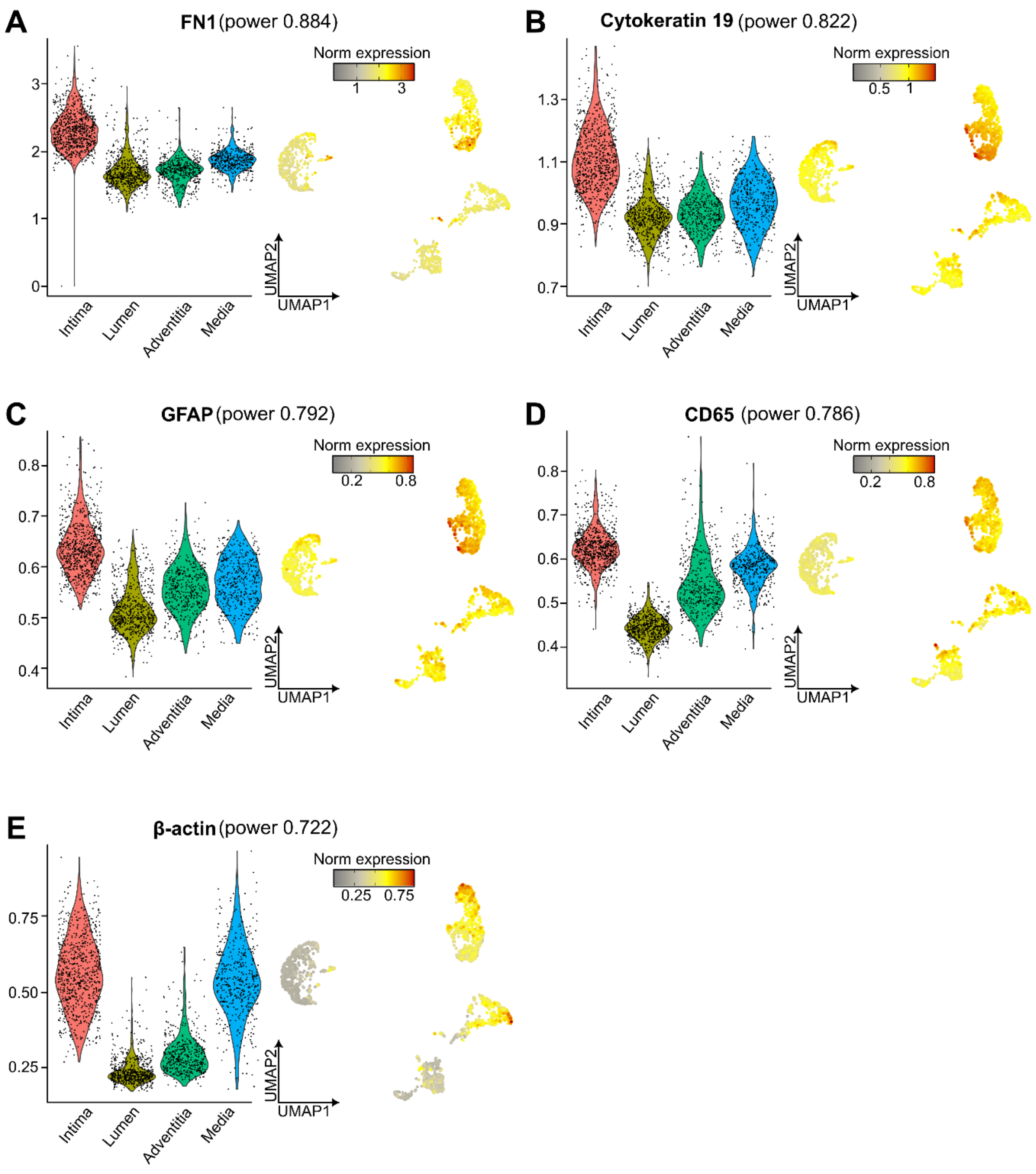


**Supplementary Figure 2. Normalized expression of marker proteins of cluster 0.** (**A-E**) Marker genes were determined using the FindAllMarkers function in Seurat using the receiver operating characteristic (roc) test and the top five features with highest classification power were displayed. For each marker normalized expression was shown in a violin plot per cluster and per cell using the Uniform Manifold Approximation and Projection (UMAP) projections. The top five markers were (**A**) Fibronectin1 (FN1), (**B**) Cytokeratin19, (**C**) Glial Fibrillary Acidic Protein (GFAP), (**D**) CD65 and (**E**) β-actin.

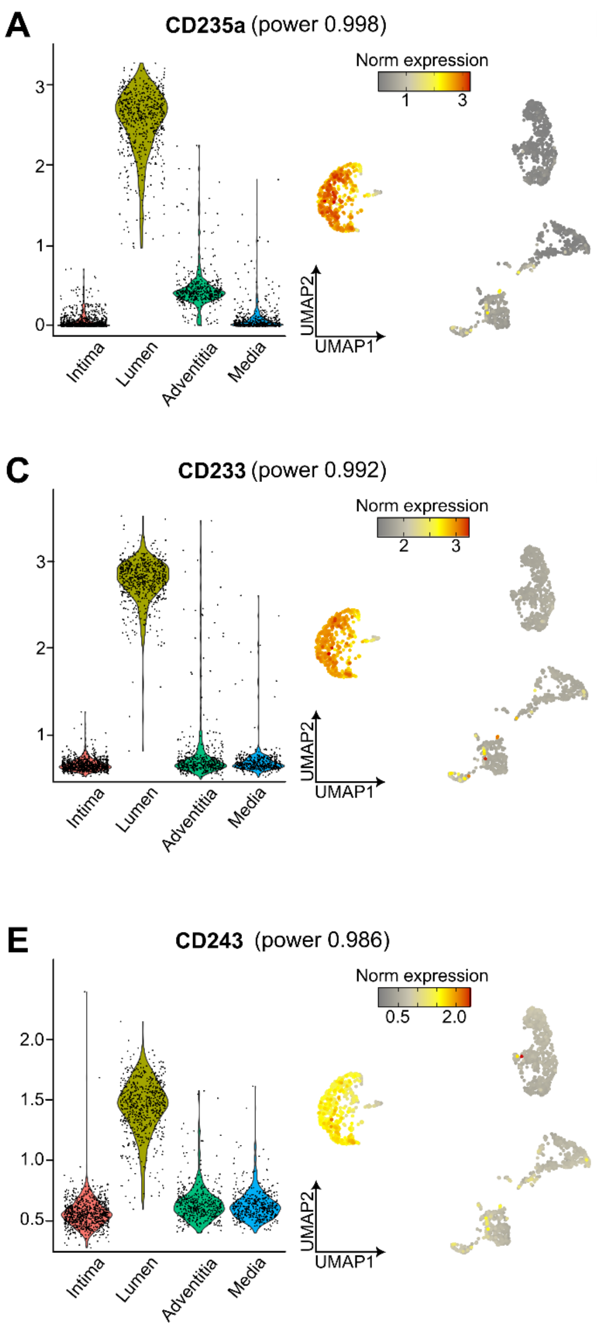


**Supplementary Figure 3. Normalized expression of marker proteins of cluster 1.** (**A-E**) Marker genes were determined using the FindAllMarkers function in Seurat using the receiver operating characteristic (roc) test and the top five features with highest classification power were displayed. For each marker normalized expression was shown in a violin plot per cluster and per cell using the Uniform Manifold Approximation and Projection (UMAP) projections. The top five markers were (**A**) CD235a, (**B**) Cytokeratin (pan-antibody), (**C**) CD233, (**D**) CD104 and (**E**) CD243.

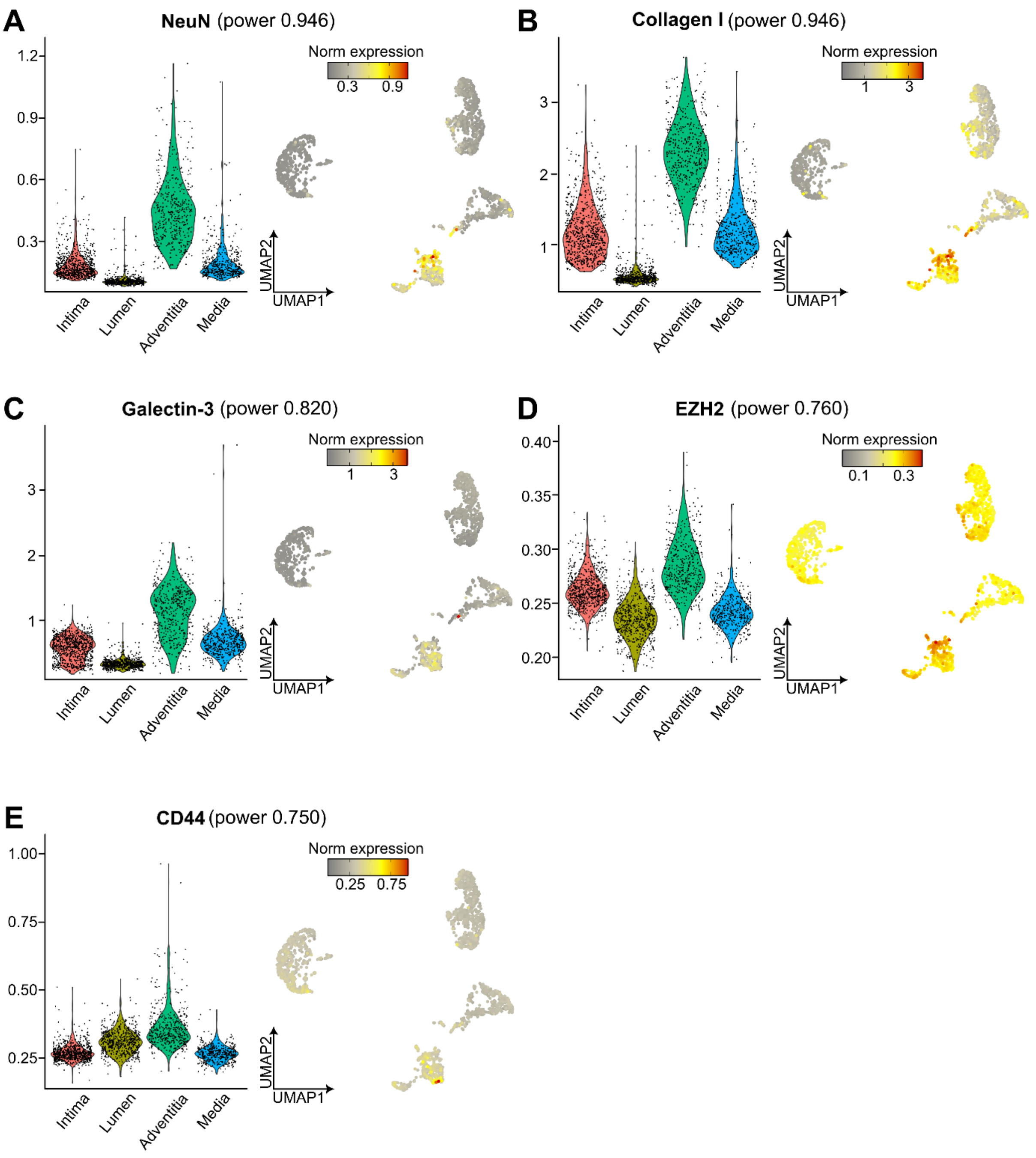


**Supplementary Figure 4. Normalized expression of marker proteins of cluster 2.** (**A-E**) Marker genes were determined using the FindAllMarkers function in Seurat using the receiver operating characteristic (roc) test and the top five features with highest classification power were displayed. For each marker normalized expression was shown in a violin plot per cluster and per cell using the Uniform Manifold Approximation and Projection (UMAP) projections. The top five markers were (**A**) NeuN (RNA binding fox-1 homolog 3), (**B**) Collagen I, (**C**) Galectin-3, (**D**) Enhancer of Zeste Homolog 2 (EZH2) and (**E**) CD44.

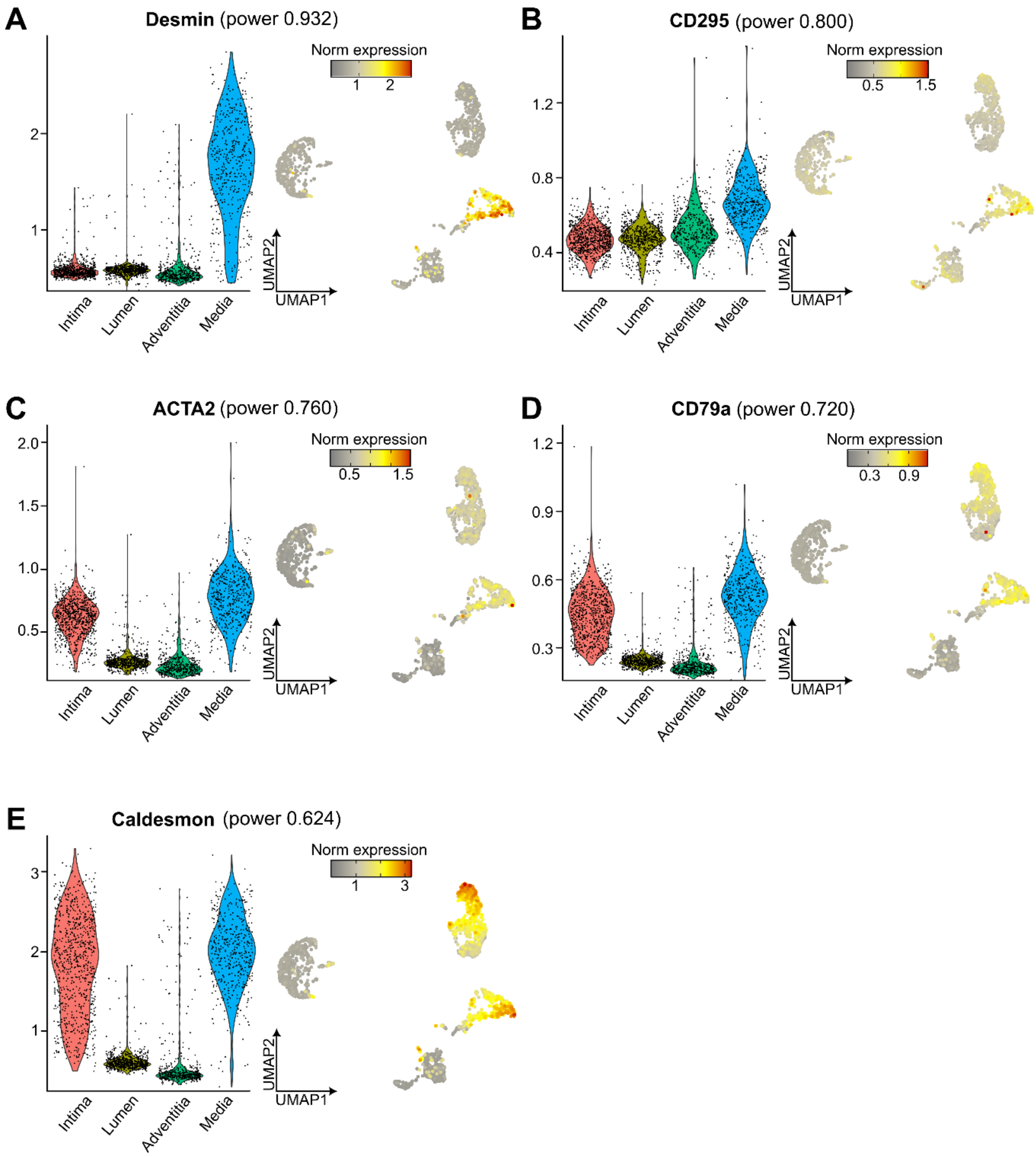


**Supplementary Figure 5. Normalized expression of marker proteins of cluster 3.** (**A-E**) Marker genes were determined using the FindAllMarkers function in Seurat using the receiver operating characteristic (roc) test and the top five features with highest classification power were displayed. For each marker normalized expression was shown in a violin plot per cluster and per cell using the Uniform Manifold Approximation and Projection (UMAP) projections. The top five markers were (**A**) Desmin (DES), (**B**) CD295, (**C**) Smoother muscle marker ACTA2, (**D**) CD79a and (**E**) Caldesmon.

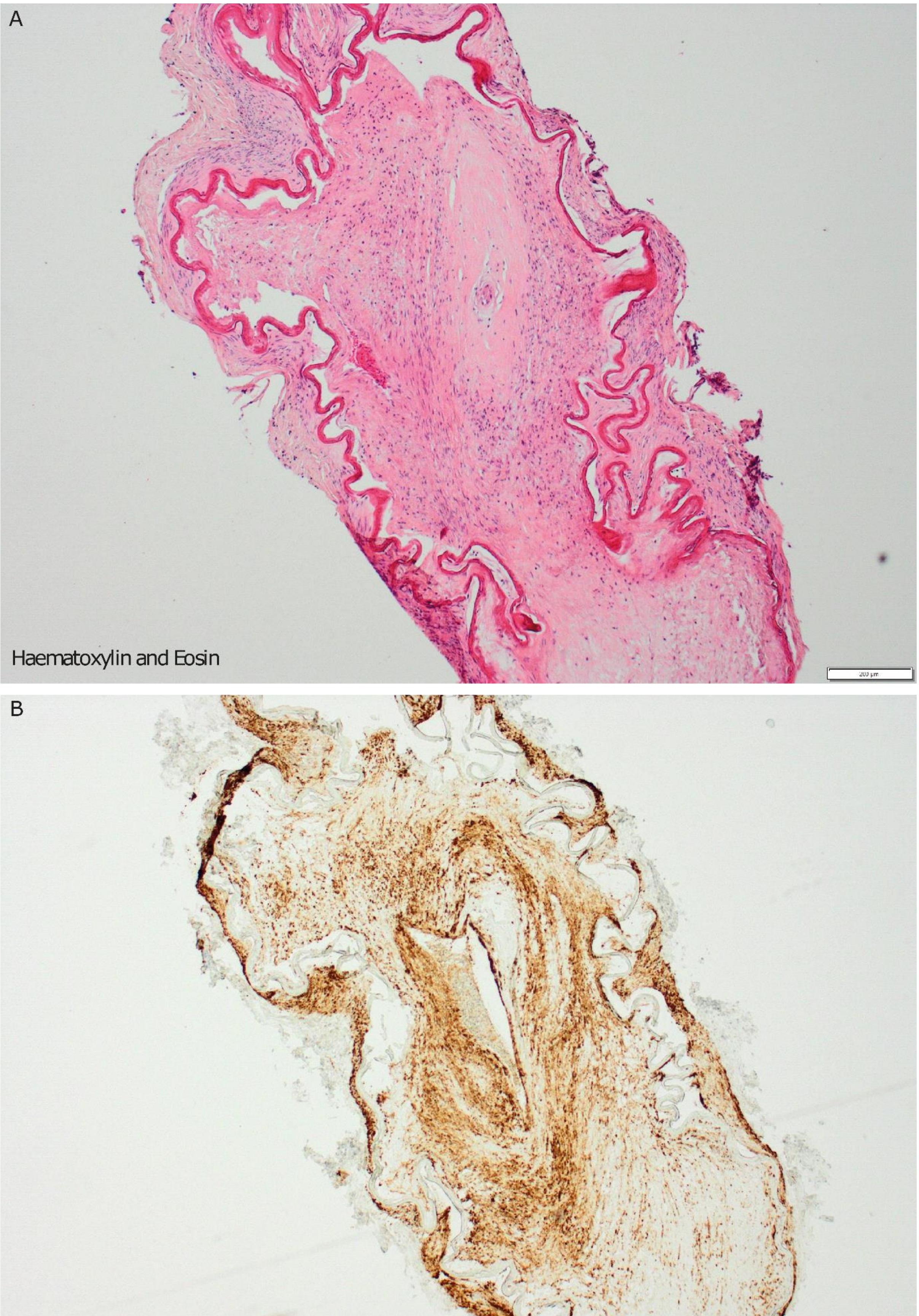
A
Haematoxylin and Eosin
200 µm
B
Smooth Muscle Actin
200 µm

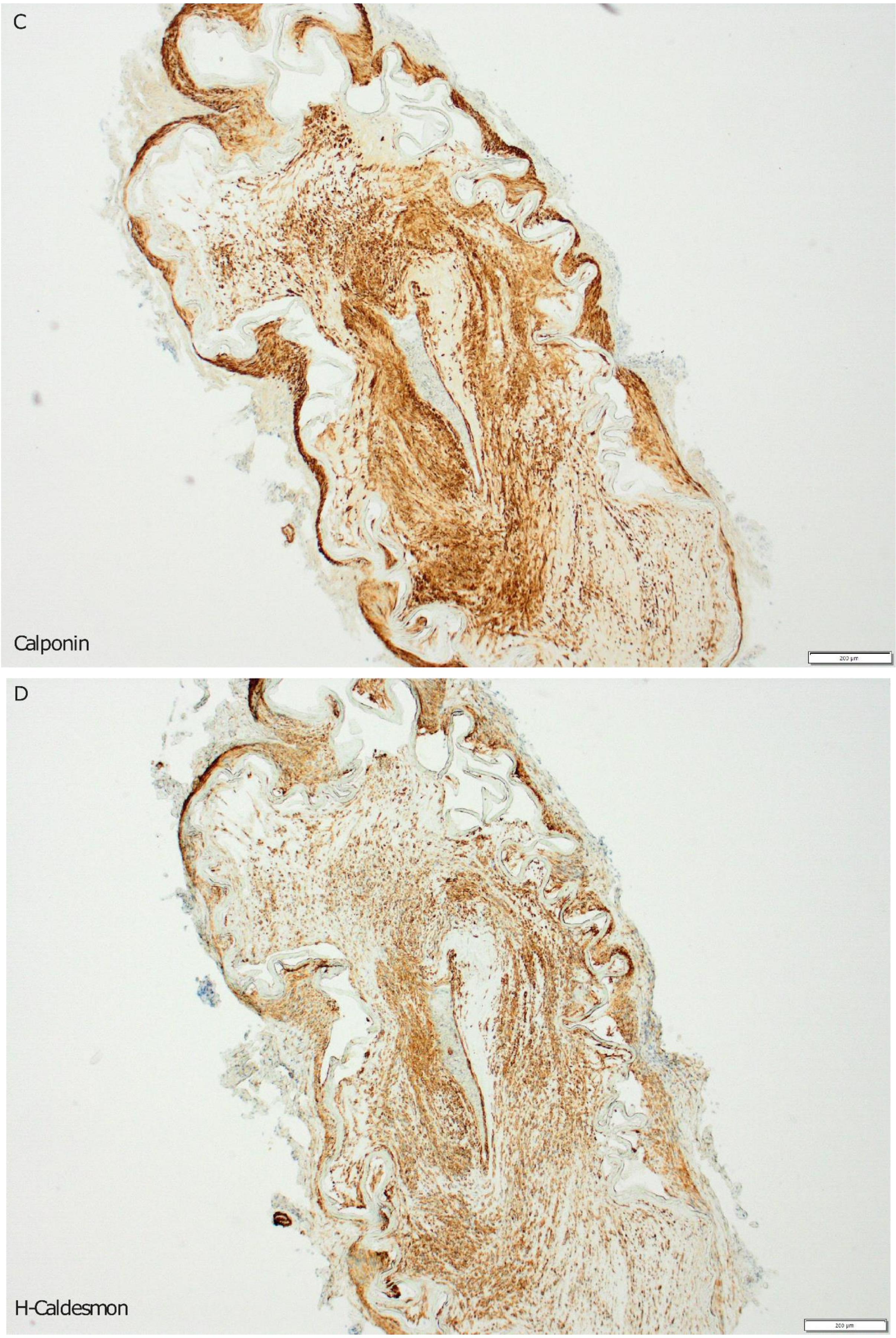
C
Calponin
D
H-Caldesmon

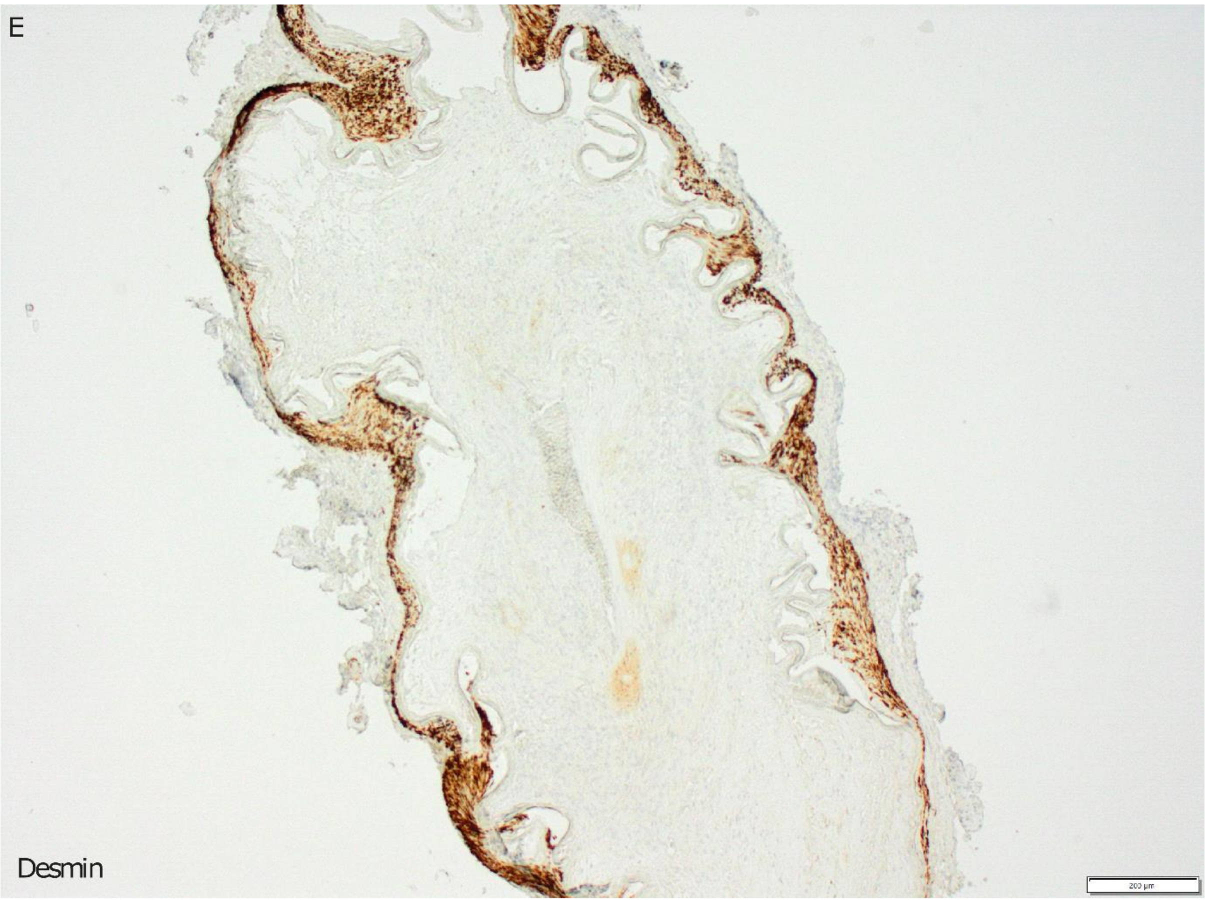
E
Desmin
200 µm

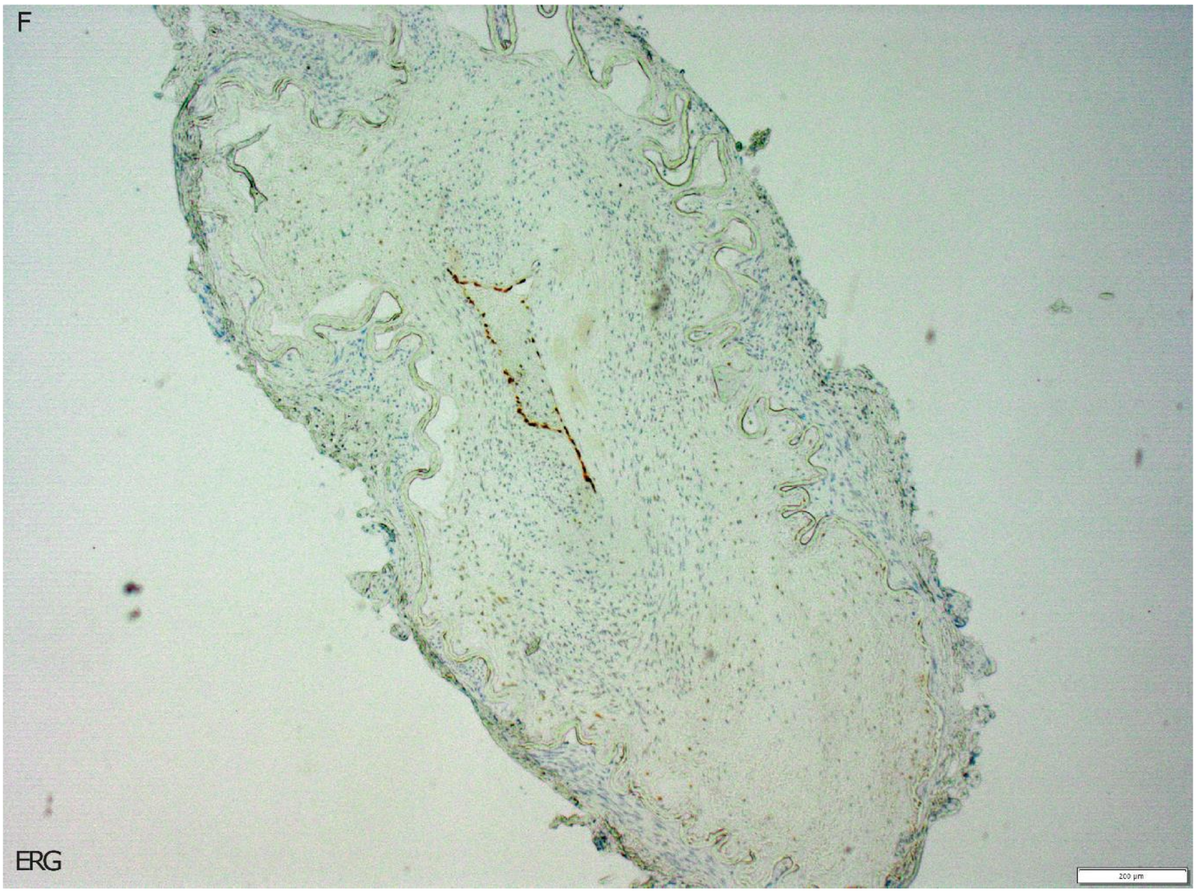
F
ERG
200 µm

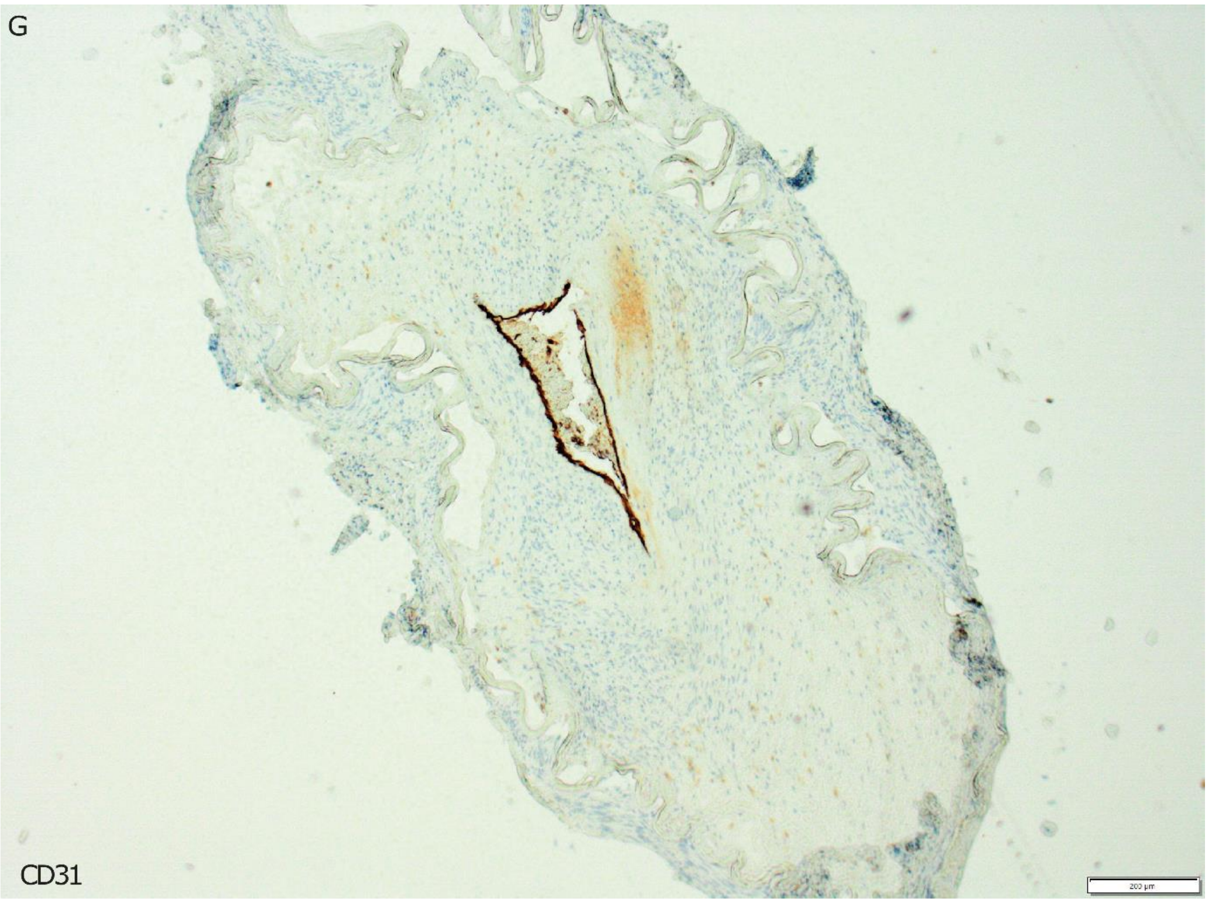
G
CD31
200 µm

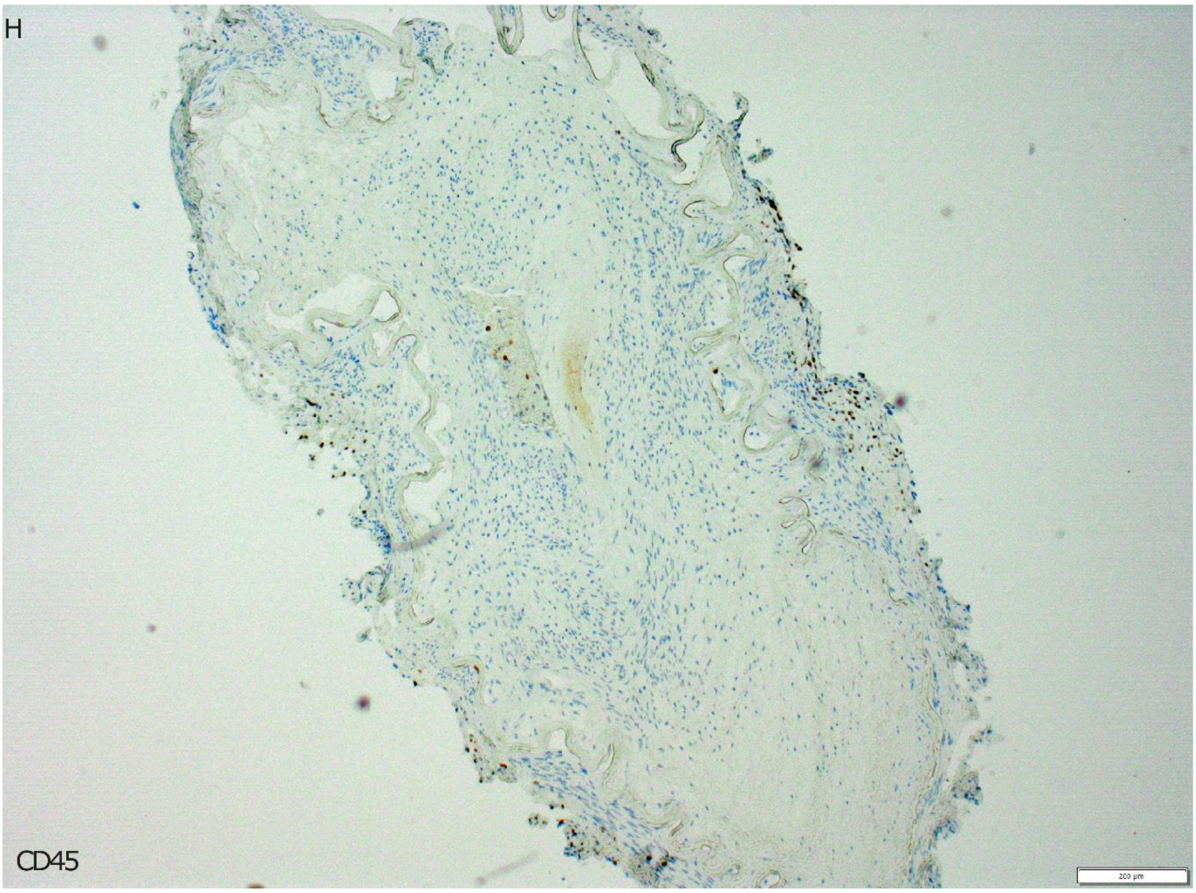
H
CD45
200 µm

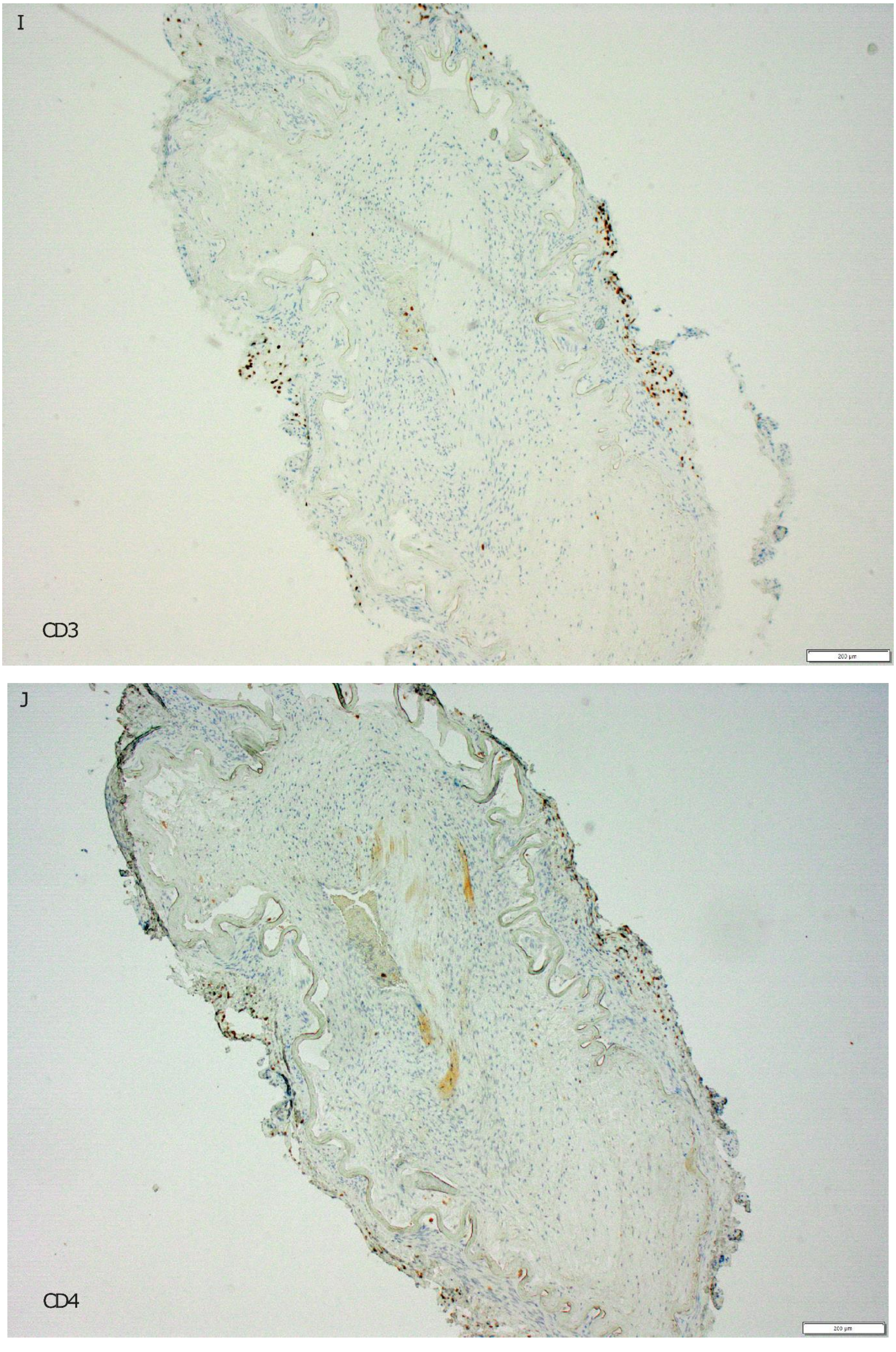
I
CD3
200 µm
J
CD4
200 µm

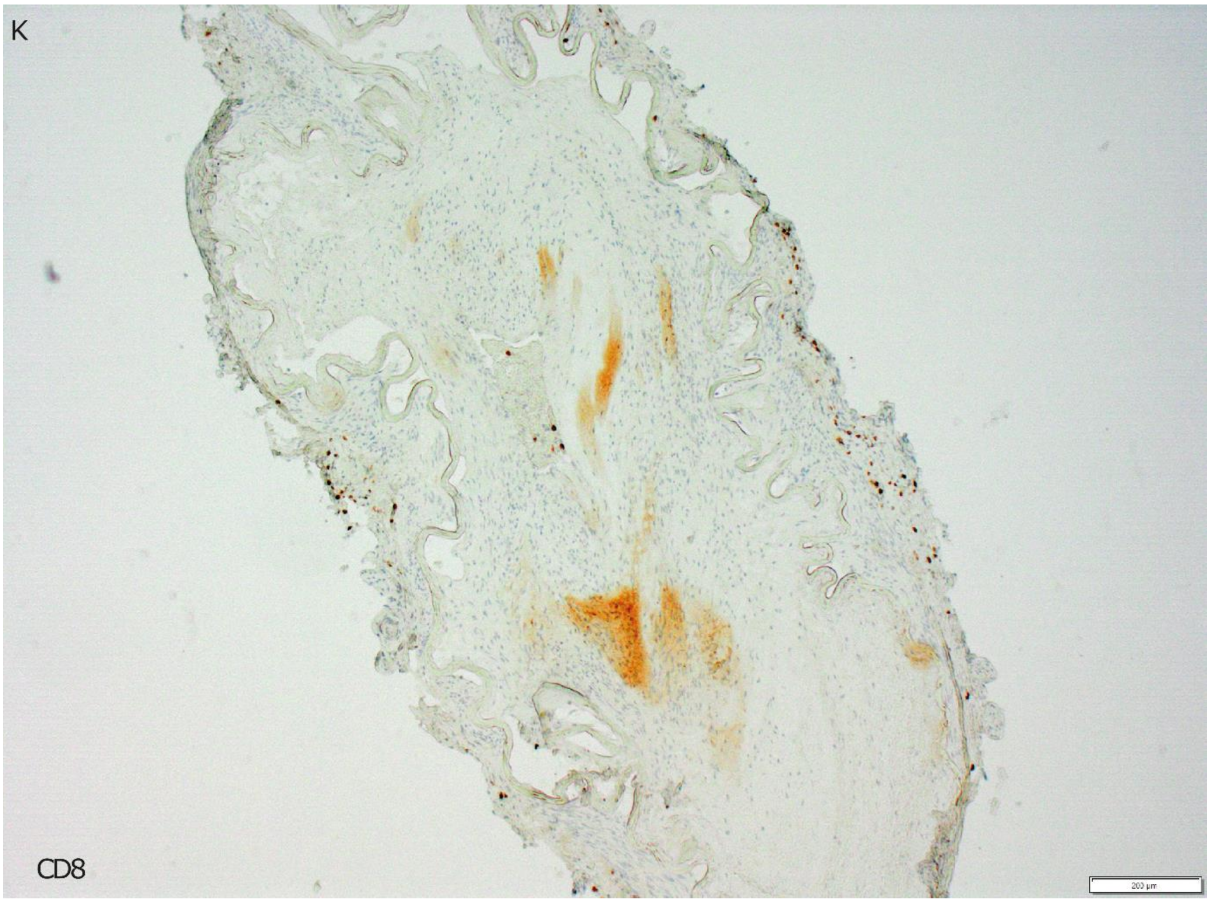
K
CD8
200 µm

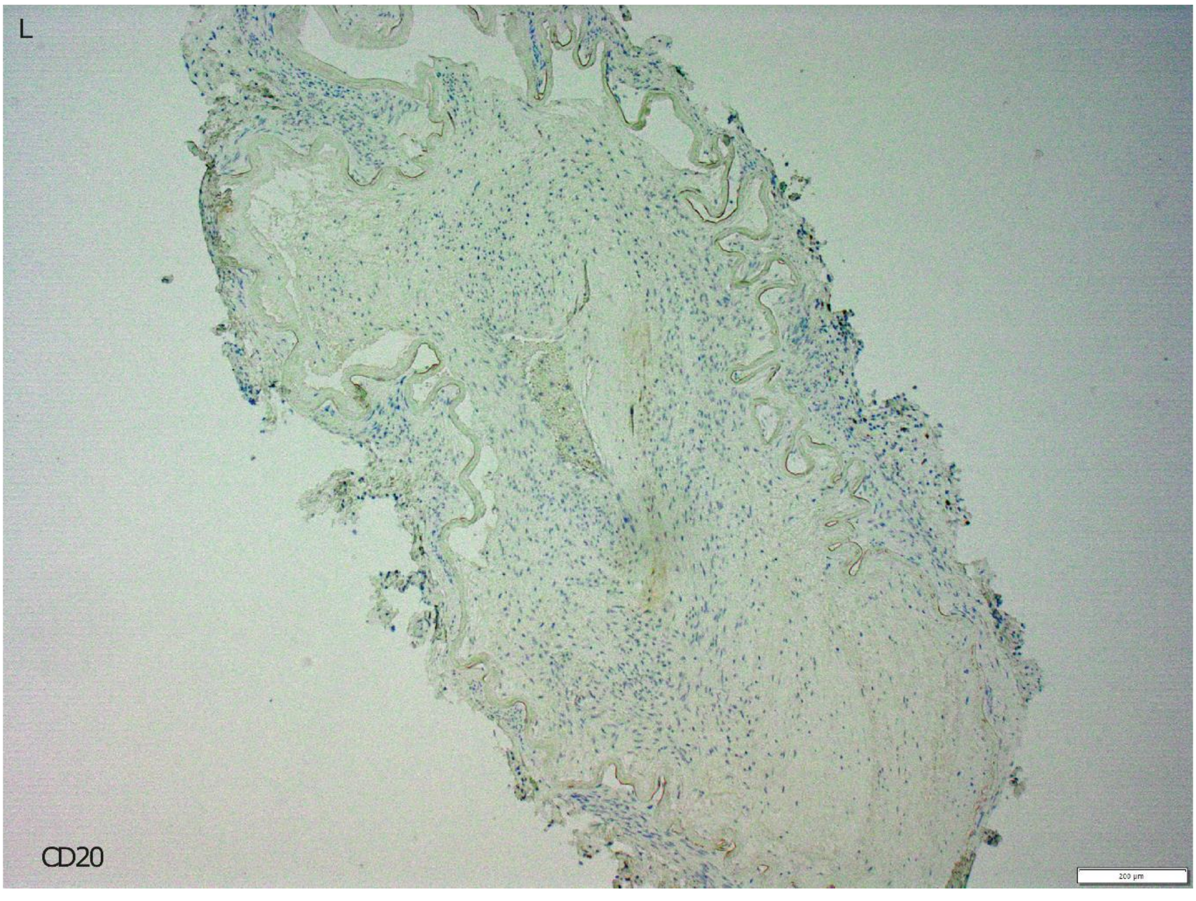
L
CD20
200 µm

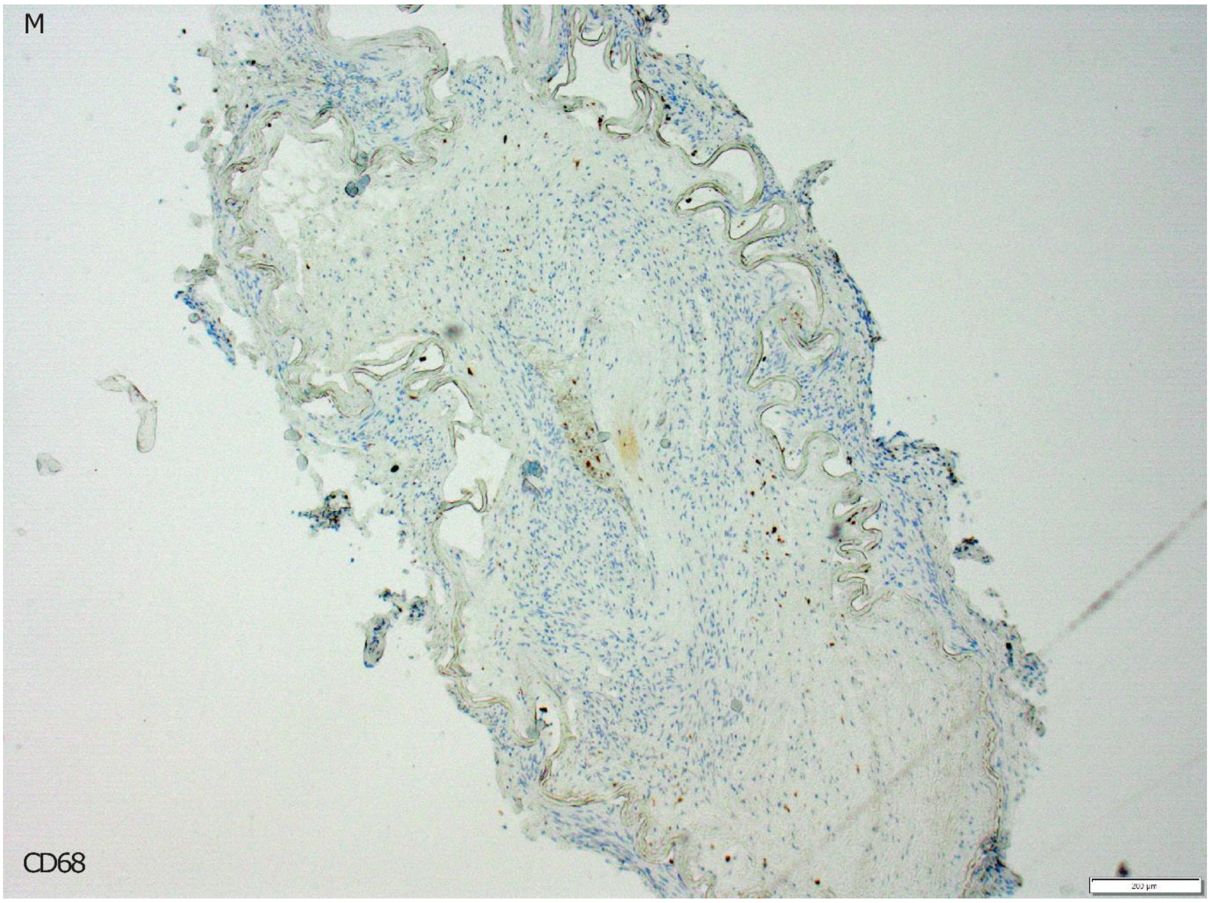


**Supplementary Figure 6. Immunohistochemistry (IHC) validation stainings. (A)** Hematoxylin and eosin (H&E) IHC validation staining was performed to assess the quality of the sample. (**B-M**) Stain for protein markers that validate cell type and immune cell annotations deducted from the spatial proteomics analysis. Stained markers included (**B**) smooth muscle actin (ACTA2), (**C**) Calponin, (**D**) H-Caldesmon, (**E**) Desmin (DES), (**F**) ERG, (**G**) CD31, (**H**) CD45, (**I**) CD3, (**J**) CD4, (**K**) CD8, (**L**) CD20 and (**M**) CD68. The white scale bar indicates 200 µm.

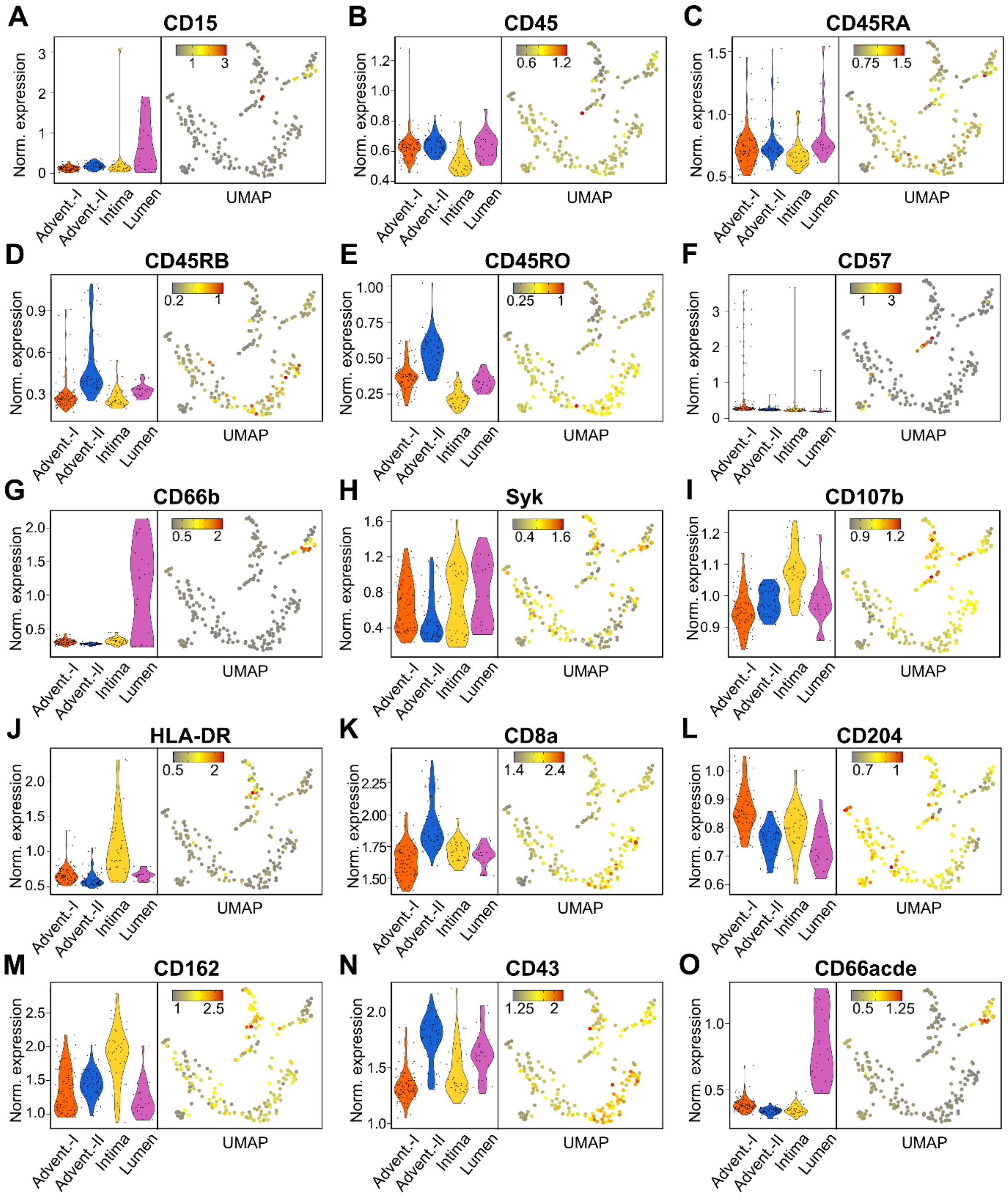


**Supplementary Figure 7. Normalized expression of immune cell related markers.** (**A-O**) Marker proteins included the 10 genes used for whole-cell segmentation of immune cells and additional proteins characteristic for immune cell subtypes. For each marker normalized expression was shown in a violin plot per cluster and per cell using the Uniform Manifold Approximation and Projection (UMAP) projections.

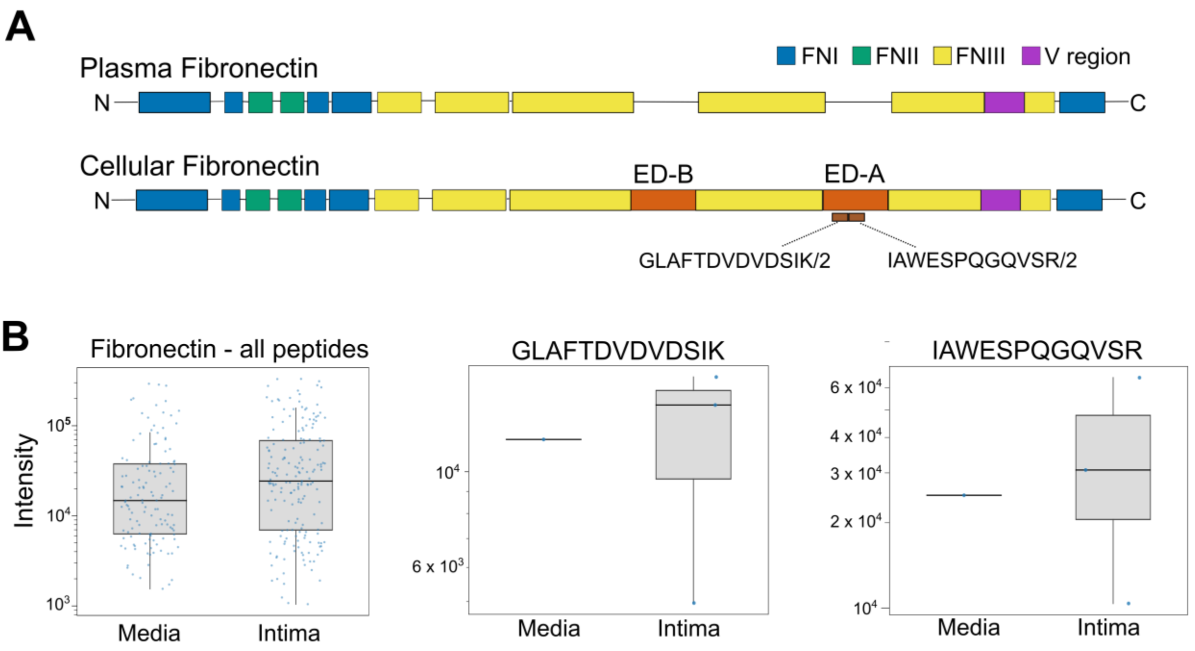


**Supplementary Figure 8. Detection of cFN1 in intimal and medial artery tissue layers.** (**A**) Schematic representation of the domain structure of plasma and cellular fibronectin (FN). Regions with FN module type I, II, and III were indicated by blue, green and yellow rectangles, respectively, while the variable region in purple. (**B**) Box plots showing the log2 peptide intensity of all detected FN1 peptides (*Left*) the GLAFTDVDVDSIK peptide (middle) and the IAWESPQGQVSR peptide (*Right*) specific for the FN1-EDA domain, for intimal and medial tissue.

#### Supplementary tables

**Supplementary Table 1: Antibodies used for immunofluorescence staining in MACSima™.**

| **Custom antibodies** | | | |
|---|---|---|---|
| Protein | Antibody name | Catalog number | Supplier |
| SG15 | ISG15 Antibody (F-9) | sc-166755 | Santa Cruz |
| RNF213 | RNF213 Polyclonal Antibody | PA5-51902 | Invitrogen |
| MMP2 | MMP2 Polyclonal Antibody, FITC conjugated | bs-0412R-FITC | Bioss Antibodies |
| FN1 | Fibronectin (1B9) Monoclonal Antibody , FITC Conjugated | bsm-33143M-FITC | Bioss Antibodies |
| IFN-γ | IFN-γ Antibody, anti-human, REAdye_lease™ | 130-125-275 | Miltenyi biotech |
| ACTA2 | alpha-Smooth Muscle Actin Antibody | NB300-978 | Novus biologicals |

| **REAscreen™ MAX Kit, human, FFPE, version 02** | | | |
|---|---|---|---|
| Antigen | Clone | Fluorochrome | Antibody type |
| Actin (Smooth Muscle) | REAL650 | FITC | REAdye_lease™ |
| AKT Pan (PKB) | REA676 | PE | REAfinity™ |
| Amyloid A | REA931 | FITC | REAfinity |
| AN2 | REA989 | PE | REAfinity |
| Annexin I | REA1122 | FITC | REAfinity |
| BATF | REA486 | FITC | REAfinity |

| | | | |
|---|---|---|---|
| Bcl-10 | REA1140 | PE | REAfinity |
| Bcl-2 | REA872 | FITC | REAfinity |
| BMI-1 | REA438 | FITC | REAfinity |
| CA-125 | REAL909 | PE | REAdye_lease |
| Caldesmon | REA1120 | PE | REAfinity |
| Calponin | REAL635 | PE | REAdye_lease |
| Catalase | REAL553 | PE | REAdye_lease |
| CD100 | REA316 | FITC | REAfinity |
| CD104 (Integrin β4) | REA236 | FITC | REAfinity |
| CD107a (LAMP-1) | REAL653 | APC | REAdye_lease |
| CD107b | REA1073 | FITC | REAfinity |
| CD117 | REA787 | PE | REAfinity |
| CD123 | REA918 | FITC | REAfinity |
| CD13 | REAL771 | PE | REAdye_lease |
| CD134 (OX40) | ACT35 | PE | Hybridoma |
| CD138 | REA929 | FITC | REAfinity |
| CD147 | REA282 | FITC | REAfinity |
| CD15 | VIMC6 | FITC | Hybridoma |
| CD158i (KIR2DS4) | JJC11.6 | PE | Hybridoma |
| CD162 | REA319 | FITC | REAfinity |
| CD181 (CXCR1) | REA958 | FITC | REAfinity |
| CD182 (CXCR2) | REA208 | PE | REAfinity |
| CD183 (CXCR3) | REAL756 | FITC | REAdye_lease |
| CD195 (CCR5) | REA245 | PE | REAfinity |
| CD196 (CCR6) | REA190 | PE | REAfinity |
| CD2 | REA1130 | FITC | REAfinity |
| CD20 Cytoplasmic | REAL649 | APC | REAdye_lease |
| CD202b (TIE-2) | REA198 | APC | REAfinity |
| CD204 | REA460 | PE | REAfinity |
| CD209 (DC-SIGN) | REAL690 | APC | REAdye_lease |
| CD223 | REA351 | PE | REAfinity |
| CD226 (DNAM-1) | REA1040 | FITC | REAfinity |
| CD233 | REA368 | FITC | REAfinity |
| CD234 (DARC) | REA376 | PE | REAfinity |
| CD235a (Glycophorin A) | REAL490 | FITC | REAdye_lease |
| CD239 (BCAM) | REA276 | PE | REAfinity |
| CD240DCE | REA327 | PE | REAfinity |
| CD243 (ABCB1) | REA495 | PE | REAfinity |
| CD268 | REA1115 | FITC | REAfinity |
| CD271 (LNGFR) | REAL709 | APC | REAlease® |
| CD279 (PD1) | REA1165 | PE | REAfinity |
| CD295 (LEPR) | REA361 | APC | REAfinity |
| CD2AP | REA1147 | PE | REAfinity |
| CD3 | REA1151 | APC | REAfinity |
| CD305 (LAIR-1) | REA447 | PE | REAfinity |
| CD317 (BST2) | REA202 | FITC | REAfinity |
| CD334 (FGFR4) | 4FR6D3 | FITC | Hybridoma |
| CD43 | REA833 | FITC | REAfinity |
| CD44 | REA690 | PE | REAfinity |
| CD45 | 5B1 | PE | Hybridoma |
| CD45RA | REAL164 | APC | REAlease |
| CD45RB | REA119 | FITC | REAfinity |
| CD45RO | REA611 | PE | REAfinity |
| CD5 | REAL760 | PE | REAdye_lease |
| CD52 | REA164 | FITC | REAfinity |
| CD57 | REA769 | PE | REAfinity |
| CD65 | REA1128 | PE | REAfinity |
| CD66 (CEA) | REA1158 | FITC | REAfinity |

| | | | |
|---|---|---|---|
| CD66abce | REA1230 | PE | REAfinity |
| CD66acde | REA428 | PE | REAfinity |
| CD66ace | REA889 | FITC | REAfinity |
| CD66b | REA306 | PE | REAfinity |
| CD66c | REA414 | FITC | REAfinity |
| CD66e | REA876 | PE | REAfinity |
| CD79a | REA1168 | PE | REAfinity |
| CD88 (C5AR) | REA1213 | PE | REAfinity |
| CD8a | REA1024 | FITC | REAfinity |
| CD90 | REAL677 | PE | REAdye_lease |
| CD99 | REAL687 | FITC | REAlease |
| CLA | REA1101 | PE | REAfinity |
| Collagen I | REAL958 | APC | REAdye_lease |
| Cytokeratin | REAL648 | APC | REAdye_lease |
| Cytokeratin 10 | REA933 | FITC | REAfinity |
| Cytokeratin 10/13 | REA1138 | FITC | REAfinity |
| Cytokeratin 14 | REAL831 | FITC | REAdye_lease |
| Cytokeratin 19 | REAL822 | PE | REAdye_lease |
| Cytokeratin 20 | REAL753 | APC | REAdye_lease |
| Cytokeratin 5/6 | REAL538 | PE | REAdye_lease |
| Cytokeratin 5/6/8/17/19 | REA1141 | FITC | REAfinity |
| Cytokeratin 7 | REA935 | FITC | REAfinity |
| Cytokeratin 8 | REA885 | FITC | REAfinity |
| Cytokeratin HMW | REAL645 | FITC | REAdye_lease |
| Dectin-1 | REA515 | PE | REAfinity |
| Desmin | REA1134 | FITC | REAfinity |
| Dnmt3b | REA565 | APC | REAfinity |
| EZH2 | REA907 | PE | REAfinity |
| FcεRIα | REA758 | FITC | REAfinity |
| Galectin-3 | REA1072 | PE | REAfinity |
| Galectin-9 | REA435 | FITC | REAfinity |
| GCDFP-15 | REAL828 | FITC | REAdye_lease |
| GFAP | REAL735 | PE | REAdye_lease |
| Hepatocyte | REA1088 | FITC | REAfinity |
| HLA-ABC | REAL597 | APC | REAdye_lease |
| HLA-DR | REAL550 | FITC | REAdye_lease |
| HLA-DR, DP, DQ | REA332 | FITC | REAfinity |
| Hsp70 | REA349 | PE | REAfinity |
| IgD | REA740 | PE | REAfinity |
| IRF-7 | REA521 | FITC | REAfinity |
| IRF-7 pS477/pS479 | REA310 | PE | REAfinity |
| Jak1 | REA700 | PE | REAfinity |
| JNK2 (MAPK9) | REA1153 | FITC | REAfinity |
| Ki-67 | REA1123 | FITC | REAfinity |
| LRP-4 | REA552 | APC | REAfinity |
| MART-1 | REA1093 | FITC | REAfinity |
| Mast Cell Tryptase | REAL798 | FITC | REAdye_lease |
| Melanocyte PMEL | REA927 | PE | REAfinity |
| Metallothionein | REA1139 | FITC | REAfinity |
| MLC2v | REA401 | PE | REAfinity |
| Myogenin | REA930 | FITC | REAfinity |
| Myosin Smooth Muscle | REA1107 | FITC | REAfinity |
| NeuN | REA1131 | PE | REAfinity |
| p120 Catenin pS879 | REA548 | PE | REAfinity |
| p53 | REA1132 | FITC | REAfinity |
| PAX-5 (BSAP) | REA140 | PE | REAfinity |
| PCNA | REA858 | FITC | REAfinity |
| PI3K p85 α | REAL589 | PE | REAdye_lease |

| | | | |
|---|---|---|---|
| PKCα | REA156 | APC | REAfinity |
| PLAP | REA1089 | FITC | REAfinity |
| Plasma Cell | REA908 | FITC | REAfinity |
| PLP | REA1155 | PE | REAfinity |
| Podoplanin | REAL468 | PE | REAlease |
| Slug | REA404 | PE | REAfinity |
| Sox2 | REA320 | FITC | REAfinity |
| SSRP1 | REA555 | PE | REAfinity |
| SUSD2 | REA795 | PE | REAfinity |
| Syk | REA111 | PE | REAfinity |
| Synaptophysin | REA1121 | FITC | REAfinity |
| TIM-3 | REAL818 | APC | REAdye_lease |
| TOM22 | REAL439 | APC | REAdye_lease |
| TTF-1 | REA1090 | FITC | REAfinity |
| Tyrosinase | REA1149 | PE | REAfinity |
| Vimentin | REAL1008 | APC | REAdye_lease |
| WT1 | REAL748 | APC | REAdye_lease |
| ZAP70 | REA814 | PE | REAfinity |
| α Tubulin | REA1136 | FITC | REAfinity |
| α-Actinin (Sarcomeric) | REA402 | PE | REAfinity |
| β-Actin | REAL571 | PE | REAdye_lease |
| β-Catenin | REA480 | FITC | REAfinity |
| β-Tubulin 3 | REA1152 | PE | REAfinity |
| γ-Actin 1 | REAL591 | FITC | REAdye_lease |

### Supplementary methods

#### Plasma proteomics

Of each sample +/- 160 µg of protein was added to a final volume of 50 µL S-Trap™ buffer containing 5% sodium dodecyl sulfate (SDS) and 50 mM triethylammonium bicarbonate (TEAB), pH 7.55. Proteins were reduced by addition of 5 mM tris(2-carboxyethyl)phosphine and incubation for 30 min at 55 ˚C and then alkylated by addition of 20 mM iodoacetamide and incubation for 15 min at room temperature in the dark. Phosphoric acid was added to a final concentration of 1.2% and subsequently samples were diluted 7-fold with binding buffer containing 90% methanol in 100 mM TEAB, pH 7.1. The samples were loaded on the 96-well S-Trap™ plate (Protifi, New York, USA), placed on top of a deepwell plate, and centrifuged for 2 min at 1,500 x g at root temperature. After protein binding, the S-Trap™ plate was washed three times by adding 400 µL binding buffer and centrifugation for 2 min at 1,500 x g at room temperature. A new deepwell receiver plate was placed below the 96-well S-Trap™ plate and 50 mM TEAB containing trypsin (1/80, w/w) was added for digestion overnight at 37 °C. By centrifugation for 2 min at 1,500 x g, peptides were eluted in three times, first with 80 µL 50 mM TEAB, then with 80 µL 0.2% formic acid (FA) in water and finally with 80 µL 0.2% FA in water/acetonitrile (ACN) (50/50, v/v). Eluted peptides were dried completely by vacuum centrifugation and stored at -20 °C until further use. For liquid chromatography-tandem mass spectrometry (LC-MS/MS) analysis, purified peptides were re-dissolved in 20 µL loading solvent A (0.1% TFA in water/ACN (98:2, v/v)) and 200 ng of each sample was loaded on an Evotip (Evosep, Denmark) according to the manufacturer's instructions. Samples were run in data-independent acquisition parallel accumulation-serial fragmentation (DIA-PASEF) mode on an Evosep One LC-system in-line connected to a timsTOF SCP mass spectrometer (Bruker). Peptides were analyzed with the 20 SPD method (72 min run length) using the endurance Evosep column (15 cm x 150 µm I.D., 1.9 µm beads, Evosep, Denmark). For elution of the peptides from the column solvent A consisted of 0.1% FA in LC-MS-grade water and solvent B consisted of 0.1% FA in ACN. Eluting peptides were measured on the timsTOF SCP in DIA-PASEF mode using a window size of 25 Da, as described before.[1]

**Laser capture microdissection proteomics profiling**

Proteins from the excised tissue samples were extracted by ultrasonication and heating in a buffer containing 5% SDS, followed by trypsin digestion and cleanup using S-Trap™ micro columns (Protifi, New York, USA), as described in detail before.[1] Resulting peptides were loaded on Evotips (Evosep, Odense, Denmark) for LC-MS/MS analysis on an Evosep One LC-system in-line connected to a timsTOF SCP mass spectrometer (Bruker) in DIA-PASEF mode. Peptides were separated on the Evosep with the 20 SPD whisper method using a 15 cm Aurora Gen3 Elite column and measured on the timsTOF SCP in DIA-PASEF mode using a window size of 25 Da, as described before.[1]

**Proteomics data analysis**

Raw DIA-PASEF data files were analyzed by library-free analysis using DIA-NN (version 1.9.0 for LCM, version 2.0.2 for plasma proteomics).[2] Firstly, a peptide precursor spectral library (peptide length 7-30) was predicted using the human UniProtKB reference proteome (20,666 proteins). Cysteine carbamidomethylation was set as a fixed modification and methionine oxidation as a variable modification, allowing one missed trypsin cleavage. All runs, of the LCM or plasma proteomics dataset, were searched against the predicted spectral library using a 15 ppm fragment and precursor ion tolerance. In case of the plasma proteomics, two samples (healthy control #8 and moyamoya disease #6) were left out due to technical considerations (suboptimal signal and identification). The outputted peptide precursor quantification matrix was used for differential protein analysis using MSqRob2 (version 1.6.1).[3] Precursor intensities were log2 transformed and subjected to sample median normalization. Precursors were required to be quantified at least twice (LCM proteomics) or three times (plasma proteomics). Afterwards, precursor intensities were summarized at the protein level, using the aggregateFeatures function, and a MSqRob model was fitted using robust linear regression with the conditions of interest specified as covariate. In case of LCM proteomics data, contrast of occlusion to VSMC tissue regions delivered nine differentially regulated proteins ($P \leq 0.01$ and fold change $\geq 2$).

For plasma proteomics, moyamoya disease plasma versus healthy, control individual identified six differentially regulated proteins (adjusted $P \leq 0.05$). The platelet contamination index was calculated using the method described by Geyer *et al.*[4] The total intensity of platelet quality markers was divided by the total intensity per sample.

#### References


1. Maia TM, Van Haver D, Dufour S, *et al.* Proteomics-Based Analysis of Laser-Capture Micro-dissected, Formalin-Fixed Paraffin-Embedded Tissue Samples. *Methods Mol Biol*. 2025;2884:333-354. doi:10.1007/978-1-0716-4298-6_20
2. Demichev V, Szyrwiel L, Yu F, *et al.* dia-PASEF data analysis using FragPipe and DIA-NN for deep proteomics of low sample amounts. *Nat Commun*. Jul 8 2022;13(1):3944. doi:10.1038/s41467-022-31492-0
3. Sticker A, Goeminne L, Martens L, Clement L. Robust Summarization and Inference in Proteome-wide Label-free Quantification. *Mol Cell Proteomics*. Jul 2020;19(7):1209-1219. doi:10.1074/mcp.RA119.001624
4. Geyer PE, Voytik E, Treit PV, *et al.* Plasma Proteome Profiling to detect and avoid sample-related biases in biomarker studies. *EMBO Mol Med*. Nov 7 2019;11(11):e10427. doi:10.15252/emmm.201910427
5. Karla AR, Pinard A, Boerio ML, *et al.* SAMHD1 compound heterozygous rare variants associated with moyamoya and mitral valve disease in the absence of other features of Aicardi-Goutieres syndrome. *Am J Med Genet A*. Apr 2024;194(4):e63486. doi:10.1002/ajmg.a.63486
